# Early Innate Immune Signatures Imprint Clinical Outcomes of *Bordetella pertussis* Challenge in a Controlled Human Infection Model

**DOI:** 10.64898/2026.06.24.26356444

**Authors:** Saeideh Jamali, May ElSherif, Kara L Redden, Hannah Burton, Nong Xu, Eric He, Christopher H. Mody, Scott Halperin, Jun Wang

## Abstract

**Background:** Despite widespread vaccination, *Bordetella pertussis* (*B. pertussis*) continues to re-emerge globally, highlighting critical gaps in our understanding of vaccine-induced protective immunity. Controlled human infection models (CHIMs) offer a powerful platform to interrogate host-pathogen interactions *in vivo*, define correlates and mechanisms of protection, and inform next-generation pertussis vaccine design.

**Methods:** This open-label, phase 1, dose-escalation CHIM trial (NCT05136599) was conducted at the Canadian Center for Vaccinology (Nova Scotia, Canada). Healthy adults aged 18-40 years with distinct infant vaccination histories (whole cell [wP] vs acellular [aP]) and low pre-existing antibody levels (anti-pertussis toxin ≤20 U/mL) were intranasally inoculated with *B. pertussis* isolate D420. Blood, serum, plasma, peripheral blood mononuclear cells (PBMCs), nasopharyngeal aspirates (NPA), and nasal washes (NW) were collected at baseline and multiple time points post-challenge. Multicolor flow cytometry assay was used to profile innate cellular immune responses, while Luminex-based assays were employed to quantify cytokines, chemokines and cytolytic molecules in NW samples and complement proteins in plasma samples. PBMC samples collected at selected timepoints were stimulated with heat-killed (HK) *B. pertussis in vitro* for assessing natural killer (NK) cell activation, effector function and maturation. Data from 59 participants receiving 10⁶ - 10^8^ colony-forming units (CFU) were analysed according to clinical outcome (non-infected, asymptomatic, symptomatic), sex, and vaccination history.

**Findings:** Although infection and symptom development followed a dose-dependent pattern, 20.34% (12/59) of participants remained non-infected and had no evidence of seroconversion across all challenge doses, suggesting the existence of intrinsic resistance enabling spontaneous clearance of infection. Notably, 91.7% (11/12) of non-infected participants were wP-immunized whereas 69.5% (16/23) of aP-immunized participants developed clinical symptoms. Innate immune profiling revealed distinct immune signatures emerging within 1–3 days after challenge among CHIM participants with different clinical outcomes. Non-infected participants exhibited sustained expansion of circulating NK cells together with early mucosal production of granzyme A, granzyme B, IL-29 (type III interferon lambda 1), and MCP-2, connecting rapid cytotoxic and antiviral-like effector programming with spontaneous clearance. In contrast, symptomatic participants displayed robust early complement activation and mucosal production of eotaxin-2 and MIP-1δ, accompanied by broad expansion of monocytes, eosinophils, and NK cells as well as depletion of circulating neutrophils in peripheral blood. Asymptomatic individuals exhibited an intermediate phenotype characterized by early I-TAC and TRAIL production with concurrent depletion of circulating neutrophils. *In vitro* assays further demonstrated that *B. pertussis* directly induced NK cell activation and degranulation, promoting production of granzymes, perforin, and IFN-γ together with CD16 upregulation. Importantly, NK-cell subset mapping revealed a hierarchical pattern linking NK-cell maturation states to clinical outcomes. Non-infected participants were enriched for adaptive/memory-like NK cells and highly cytotoxic NK1C subsets, whereas symptomatic participants exhibited marked attenuation of NK-cell maturation and expansion of immature NK2 subset.

**Interpretation:** Our results demonstrate that distinct early innate immune programs are associated with divergent clinical trajectories following *B. pertussis* challenge. Robust type 1 memory-like NK-cell responses likely serve as an effective first line of defense, promoting spontaneous bacterial clearance through direct cytotoxicity and antibody-dependent cellular cytotoxicity (ADCC) in non-infected participants. In contrast, increased production of type 2 inflammatory mediators, together with expansion of immature NK2 subsets, is associated with a less favorable immune environment for bacterial control. This state is accompanied by pronounced complement activation and broader engagement of innate and adaptive immune responses. Collectively, these findings reveal a previously underappreciated role for memory-like cytotoxic NK-cell responses in mediating sterilizing immunity and identify NK cell–mediated protective mechanisms as promising targets for the rational design of next-generation pertussis vaccines. Our results further highlight adaptive/memory-like cytotoxic NK cells as promising candidates for vaccine-induced immunological correlates of protection.

**Funding:** Centers for Disease Control and Prevention (CDC), National Institutes of Health (NIH), Canadian Institute of Health Research (CIHR), IWK Health Center

**RESEARCH IN CONTEXT:** *Evidence before this study:* Despite high pertussis vaccine coverage worldwide, whooping cough remains a persistent public health challenge, with recurrent outbreaks occurring in many countries. Although adaptive humoral and cellular immune responses induced by wP and aP vaccines have been extensively characterized and are widely used as immunological correlates of protection, the immune mechanisms responsible for sterile immunity that prevents infection and enables spontaneous clearance of *B. pertussis* remain poorly understood. In particular, the contribution of innate immune responses and NK cells to protection against *B. pertussis* has not been systematically investigated in humans.

*Added value of this study:* Using a CHIM study, we found that approximately 20% of participants remained uninfected following *B. pertussis* challenge, indicating the presence of effective host defense mechanisms that enable spontaneous pathogen clearance. Resistance to *B. pertussis* infection was largely associated with a history of wP vaccination and was characterized by selective sustained expansion of circulating CD56^dim^ cytotoxic NK cells, early mucosal production of cytotoxic mediators, and enrichment of adaptive/memory-like NK cells and highly cytotoxic NK-cell subsets with enhanced degranulation and ADCC capacity. These findings identify NK-cell-mediated cytotoxic effector functions as a previously underappreciated first line of defense against *B. pertussis* and suggest a potential novel immunological correlate of vaccine-induced sterilizing immunity.

*Implications of all the available evidence:* The contribution of NK cells to sterile protective immunity against *B. pertussis* has likely been underestimated. Our findings implicate NK cell-mediated cytotoxicity and memory-like NK-cell responses as important determinants of resistance to infection and spontaneous bacterial clearance. These discoveries support the incorporation of NK cell-targeted approaches into the development of next-generation pertussis vaccines and highlight the need to evaluate NK-cell functionality as a vaccine-induced immunological correlate of protection.

**GRAPHIC ABSTRACT:** 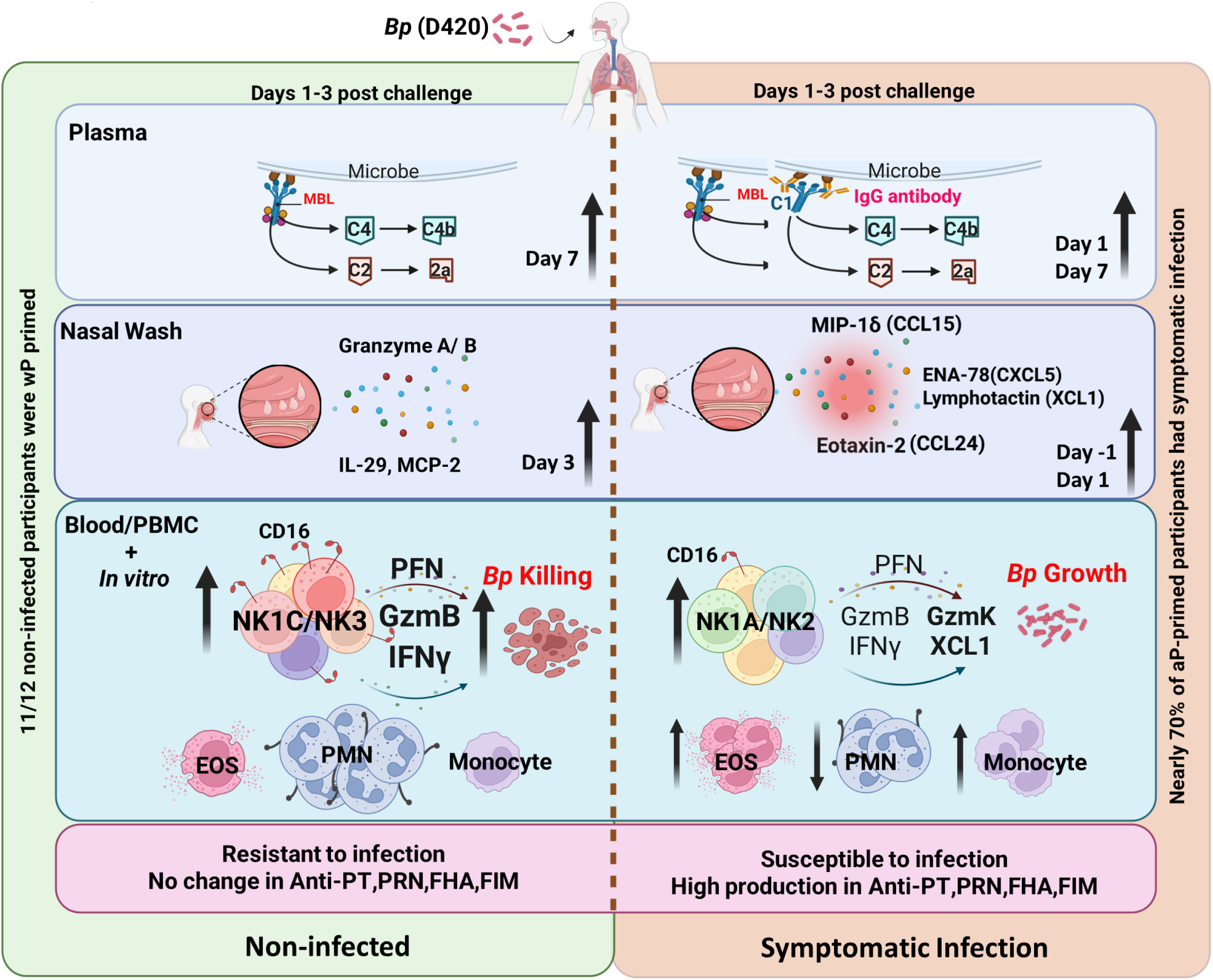

Early innate immune signatures imprint host susceptibility and clinical outcomes to *B. pertussis* challenge.

## INTRODUCTION

Pertussis (whooping cough) is a highly contagious respiratory disease caused by the Gram-negative bacterium *Bordetella pertussis* (*B. pertussis*). Following colonization of the nasopharyngeal mucosa, the pathogen proliferates locally and disrupts respiratory epithelial integrity through the coordinated production of multiple virulence factors and toxins. This results in epithelial damage, impaired mucociliary clearance, and robust activation of innate and adaptive immune responses, ultimately leading to the characteristic paroxysmal cough and clinical manifestations that can persist for weeks to months.^1^ Although routine childhood immunization programs have been in place for more than six decades, pertussis remains a significant cause of vaccine-preventable disease globally, responsible for approximately 161,000 childhood deaths annually.^2, 3^ The continued circulation of *B. pertussis* highlights limitations in the durability and effectiveness of current pertussis vaccines and underscores the need for next-generation vaccines and vaccination strategies that induce sterilizing immunity, prevent nasopharyngeal colonization, and promote rapid pathogen clearance to interrupt transmission.

The resurgence of pertussis is likely driven by multiple interacting factors, including the transition from whole-cell pertussis (wP) to acellular pertussis (aP) vaccines and associated waning immunity, pathogen evolution, and increased case detection due to improved molecular diagnostics and heightened surveillance.^2^ However, fundamental limitations of aP vaccines in inducing protective immunity at the nasal mucosa may play a central role.^4, 5^ Using diverse animal models, including mice and baboons, investigators have extensively characterized immune responses elicited by wP and aP vaccination, as well as by natural infections. Collectively, these studies indicate that aP vaccines have several fundamental limitations compared with wP vaccines. First, aP-induced protection is relatively short-lived.^6^ Pertussis outbreaks among vaccinated children and adolescents have been attributed in part to waning immunity, reflected by declining circulating antibody titers following aP immunization.^7, 8^ Second, aP vaccination induces a predominantly Th2-skewed immune response and promotes IgG4 production,^4, 9, 10^ whereas wP vaccination or natural infection elicits robust Th1/Th17 responses, facilitates the production of opsonising IgG subclasses (IgG1 and IgG3), and enhances neutrophil recruitment and activation, thereby supporting more effective bacterial clearance.^9, 11, 12^ The four human IgG subclasses (IgG1-IgG4) differ substantially in effector function, particularly in their capacity to engage Fcγ receptor-expressing cells, mediate opsonophagocytosis and ADCC activity, and activate the classical complement pathway^13^. IgG1 and IgG3 are potent activators of the classical complement pathway, whereas IgG2 is a weak activator, and IgG4 is generally considered inactive—or even functionally inhibitory—in this context.^14^ Accordingly, IgG-mediated opsonization and complement activation have emerged as central vaccine-induced protective mechanisms against *B. pertussis*.^2, 15, 16^. However, it remains unclear whether these systemic IgG antibody-mediated mechanisms are sufficient to confer sterilizing immunity capable of preventing nasopharyngeal colonization and infection in humans.

Furthermore, wP vaccination induces prolonged tissue resident memory T (T_RM_) cells at the respiratory mucosa and confers robust protection through IL-17-dependent neutrophil mobilization upon *B. pertussis* challenge.^5, 17^ In sharp contrast, aP vaccination limits the generation of mucosal Th17 memory T cells, resulting in reduced protection within the nasopharyngeal compartment.^18, 19^ While these studies highlight a protective role for T_RM_ cells in host defense against *B. pertussis*, bacterial clearance in these animal studies typically requires at least two weeks following challenge.^5, 18, 19^. Therefore, it remains debatable whether IL-17-mediated neutrophil responses are sufficient to achieve true sterilizing immunity that completely prevents initial colonization.

Recent advances in immunology have introduced the concept of trained immunity, whereby innate immune cells—including NK cells, monocytes and macrophages—undergo functional reprogramming following pathogen exposure or immunization, resulting in enhanced memory-like responses upon secondary exposure.^20^ In a recent asymptomatic *B. pertussis* CHIM study by the PERISCOPE consortium, early peripheral blood cellular responses, including monocytes and NK cells, differ significantly between participants who had colonization and those who remained uncolonized.^21^ However, whether innate immune cells mediate sterilizing protective immunity against *B. pertussis* remains unclear. In the present CHIM study^22^, we specifically investigated early innate immune responses following *B. pertussis* challenge in individuals previously primed with either wP or aP vaccines. We demonstrate that distinct early innate immune signatures imprint clinical outcomes after challenge, with enhanced NK cell cytotoxic effector function and the development of memory-like NK cells strongly associated with sterilizing protection.

## METHODS

### Pertussis CHIM study design, eligibility criteria, and clinical outcomes

Healthy men and women aged 18-40 years were enrolled based on pertussis-specific eligibility criteria: no history of laboratory-confirmed pertussis; ≥5 years since last booster; ≤7 lifetime pertussis vaccine doses; baseline anti-pertussis toxin (PT) IgG ≤20 IU/mL; and no evidence of *B. pertussis* carriage by negative nasal culture and PCR. Participant demographic characteristics are presented in Supplementary Table 1 (**S-Table 1**). The protocol was approved by the IWK Health Research Ethics Board with written informed consent obtained from all participants. Among enrolled participants, 59 who received the challenge dose (10⁶–10^8^ CFU) were included in the analysis (**S-Table 2**).

The administered inoculum was confirmed by plating serial dilutions of challenge material. Infection was monitored daily using NW and NPA samples, with *B. pertussis* detected by culture and PCR targeting *ptxS1* and *IS481*. Serum antibodies to pertussis antigens (PT, FHA, PRN, FIM) were measured at Days -1, 28, 42, and 56 using ELISA. Based on predefined microbiological and clinical criteria^22^, participants were classified into three outcome groups: (1) Symptomatic infection, defined as meeting microbiological criteria (≥1 positive culture and/or ≥3 positive PCR results from ≥day 6 post-challenge to azithromycin initiation) plus ≥2 solicited symptoms, including ≥1 respiratory symptom; (2) Asymptomatic infection, meeting microbiological criteria without fulfilling symptom criteria; and (3) Non-infected, with no evidence of infection (no positive culture and <3 positive PCR results), with reported symptoms considered unrelated to pertussis. Biological samples, including NW, NPA, serum, plasma and PBMCs, were collected at different timepoints and approved for the assessment of host immune responses using a range of immunological assays.

### Assessment of Innate Immune Responses by Flow Cytometry and Luminex Assay

A multicolor flow cytometry panel was designed to longitudinally monitor innate immune cell subsets in peripheral blood. Samples were acquired on a BD Biosciences LSRFortessa, with at least 200,000 CD45⁺ events collected per sample, and data were analyzed using De Novo Software FCS Express version 7. Cytokine and chemokine levels in NW samples (Days -1, 1, and 3) were measured using a 48-plex multiplex assay, while plasma complement proteins (Days -1, 1, and 7) were quantified using human complement Luminex panels. Details of the antibodies and assay kits used in this study are provided in **S-Material and Method**.

### NK cell activation, phenotyping and subset mapping

To assess the impact of *B. pertussis* on NK cell activation and degranulation, 1 × 10⁶ YT NK cells or PBMCs, with or without monocyte depletion, were stimulated with HK *B. pertussis* at MOI of 100 for 6 or 24 hours. Anti-CD107a antibody was added at the initiation of culture to capture surface CD107a expression as a marker of NK cell degranulation. Brefeldin A and monensin were added during the final 4 hours of culture to facilitate intracellular cytokine and effector molecule detection. Following stimulation, cells were harvested and subjected to surface and intracellular staining for flow cytometric analysis. Data were analyzed using De Novo Software FCS Express version 7. NK cell subsets were visualized using t-distributed stochastic neighbor embedding (t-SNE) from representative samples (n=4 per group) and annotated based on described phenotypic characteristics.^23^ Representative genes associated with each NK cell subset were obtained from the Human Cell Atlas (https://cellatlas.io/studies/meta-nk/dataset) and visualized as a heatmap in R.

### Statistical analysis

All statistical analyses were performed using GraphPad Prism (version 10), with the specific tests indicated in the corresponding figure legends. Data normality was assessed prior to selecting appropriate tests, and statistical significance was defined as p < 0.05.

## RESULTS

### CHIM participants display heterogenous clinical outcomes following *B. pertussis* challenge

To characterize innate immune signatures elicited by *B. pertussis* challenge, we analyzed 59 participants who received challenge doses ranging from 10⁶ to 10^8^ CFU (**Fig.1a).** Although infection and symptom rates increased in a dose-dependent manner, 20.23% (12/59) of participants remain non-infected across all challenge doses (**Fig.1a** and **S-Table 2**), suggesting intrinsic resistance to *B. pertussis* infection. These individuals had no detectable bacteria as early as day 3 post-challenge and remained free of detectable bacteria throughout the study period, despite receiving challenge doses comparable to those administered to symptomatic and asymptomatic participants (**Fig.1b**, **1c**). A total of 23.73% (14/59) of participants remained clinically asymptomatic despite substantial bacterial colonization (**Fig.1a-1c**). The remaining 55.9% (33/59) developed pertussis symptoms, accompanied by a significantly increased bacterial burden at days 9 and 12 post challenge (**Fig.1b**) and a robust induction of anti-pertussis antibodies relative to baseline levels (**Fig.1d**). In contrast, non-infected participants and most asymptomatic individuals did not show seroconversion (**Fig.1c**, **1d**), suggesting that antigen exposure was insufficient in magnitude or duration to trigger detectable adaptive immune activation and antibody production. Male sex and prior wP vaccination were associated with an increased likelihood of resistance to infection, independent of inoculum dose (**Fig.1e–1h**). While baseline anti-PT IgG levels were higher in non-infected individuals than in symptomatic participants (**S-Fig.1a**), baseline antibody levels were comparable between males and females and between wP- and aP-immunized individuals **(S-Fig.1b-1c)**, suggesting that the enhanced resistance associated with male sex and wP vaccination is unlikely to be attributable to differences in pre-existing humoral immunity. Collectively, these data demonstrate heterogenous clinical trajectories following *B. pertussis* challenge in CHIM participants, with approximately 20% remaining non-infected and without measurable adaptive immune activation.

**Figure 1:**
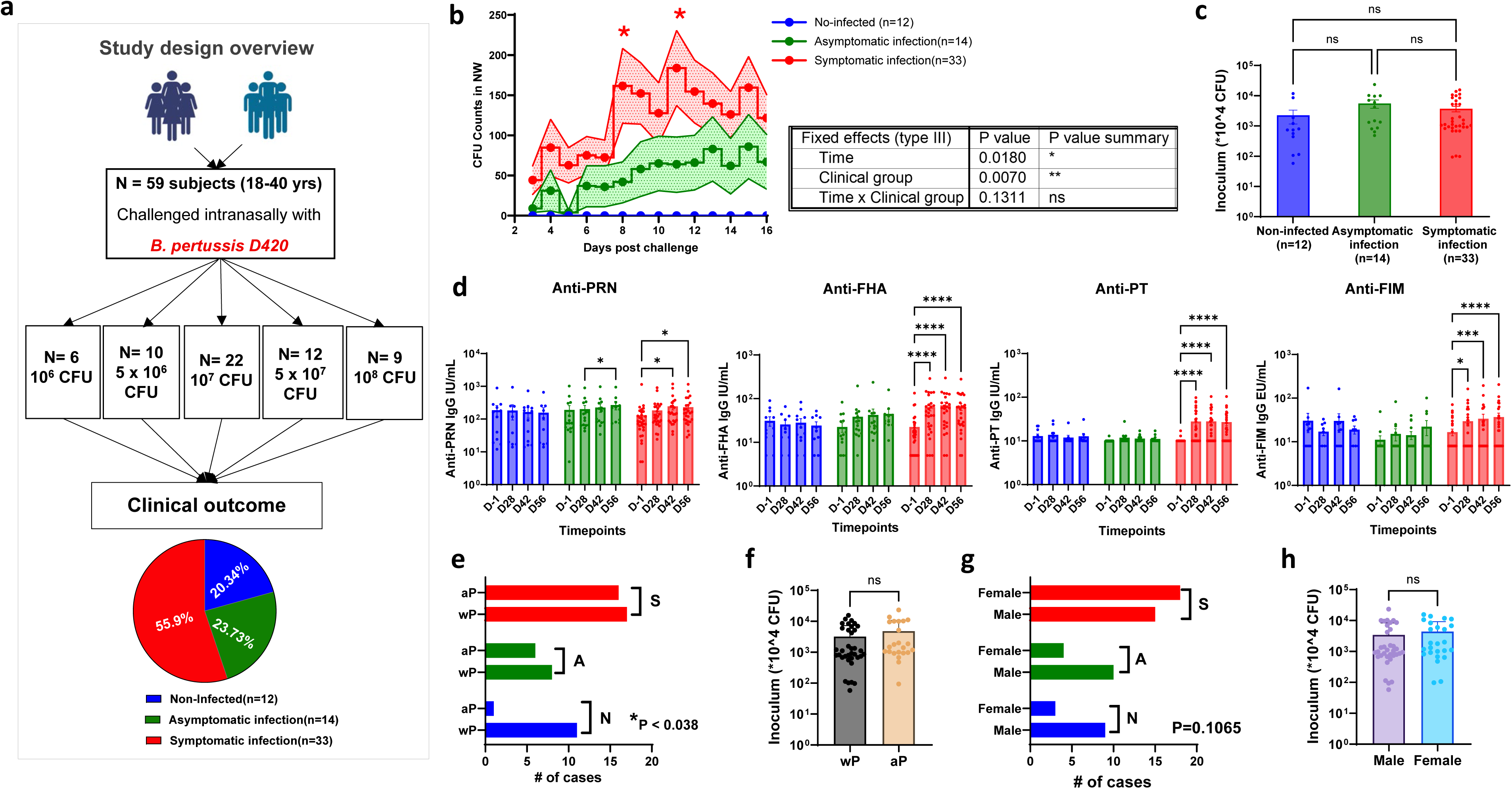
CHIM participants display distinct clinical outcomes following *B. pertussis* challenge. (a) Study overview. (b) *B. pertussis* detection in nasal wash samples by culture. (c) Confirmed bacterial inoculum doses in different groups. (d) Longitudinal serum antibody responses measured by ELISA specific for pertactin (PRN), filamentous hemagglutinin (FHA), pertussis toxin (PT), and fimbriae (FIM). (e) Distribution of participants with different clinical outcomes by childhood vaccination history.(f) Confirmed bacterial inoculum dose in aP-versus wP-immunized participants. (g) Distribution of participants with different clinical outcomes by sex. (h) Confirmed bacterial inoculum dose in male versus female participants. Data are presented as percentages or mean ± SEM. Statistical analyses: two-way ANOVA with Tukey’s post-test for panels (b) and (d), Kruskal-Wallis test for panel (c), Mann-Whitney test for panels (f) and (h), Fisher’s exact test for panels (e) and (g), Statistical significance: *p < 0.05, **p < 0.01, ***p < 0.001, ****p < 0.0001; ns: not significant. In panels (e-g), N: Non-infected; A: Asymptomatic infection; S: Symptomatic infection.

### *B. pertussis* challenge elicits different complement activation patterns across three clinical outcome groups

To understand why non-infected individuals are capable of rapidly clear bacteria within 3 days of challenge, we sought to define the early innate immune signatures associated with distinct clinical outcomes in CHIM participants. Given the central role of the complement in opsonization, membrane attack complex (MAC) formation, and innate defense against *B. pertussis,*^15, 24^ we first measured complement proteins, activated complement fragments, and regulatory factors in plasma samples collected at baseline and days 1 and 7 post challenge. While complement activation was seen in all participants by day 7 post challenge, distinct complement activation kinetics were observed across clinical outcomes (**Fig.2**). Symptomatic participants, but not non-infected or asymptomatic individuals, exhibited early and robust activation of the classical/lectin complement pathways at day 1 post challenge. This was characterized by reduced circulating levels of C1q, C2, C4, and C3, consistent with complement consumption, accompanied by increased concentrations of the activation fragments C4b and C5a, as well as elevated terminal pathway components C5 and C9 (**Fig.2a**). Notably, non-infected participants showed selective early consumption of C2, but not C1q, at day 1 post-challenge, suggesting preferential activation of the lectin pathway rather than the classical pathway. Furthermore, an early consumption of properdin was only observed in non-infected participants at day 1 post challenge (**Fig.2b**), indicating early engagement of the alternative pathway. These findings suggest that non-infected individuals mount an early complement response characterized by preferential activation of the lectin and alternative pathways, whereas symptomatic participants develop a strong and early complement activation dominated by the classical/lectin pathways.

**Figure 2.**
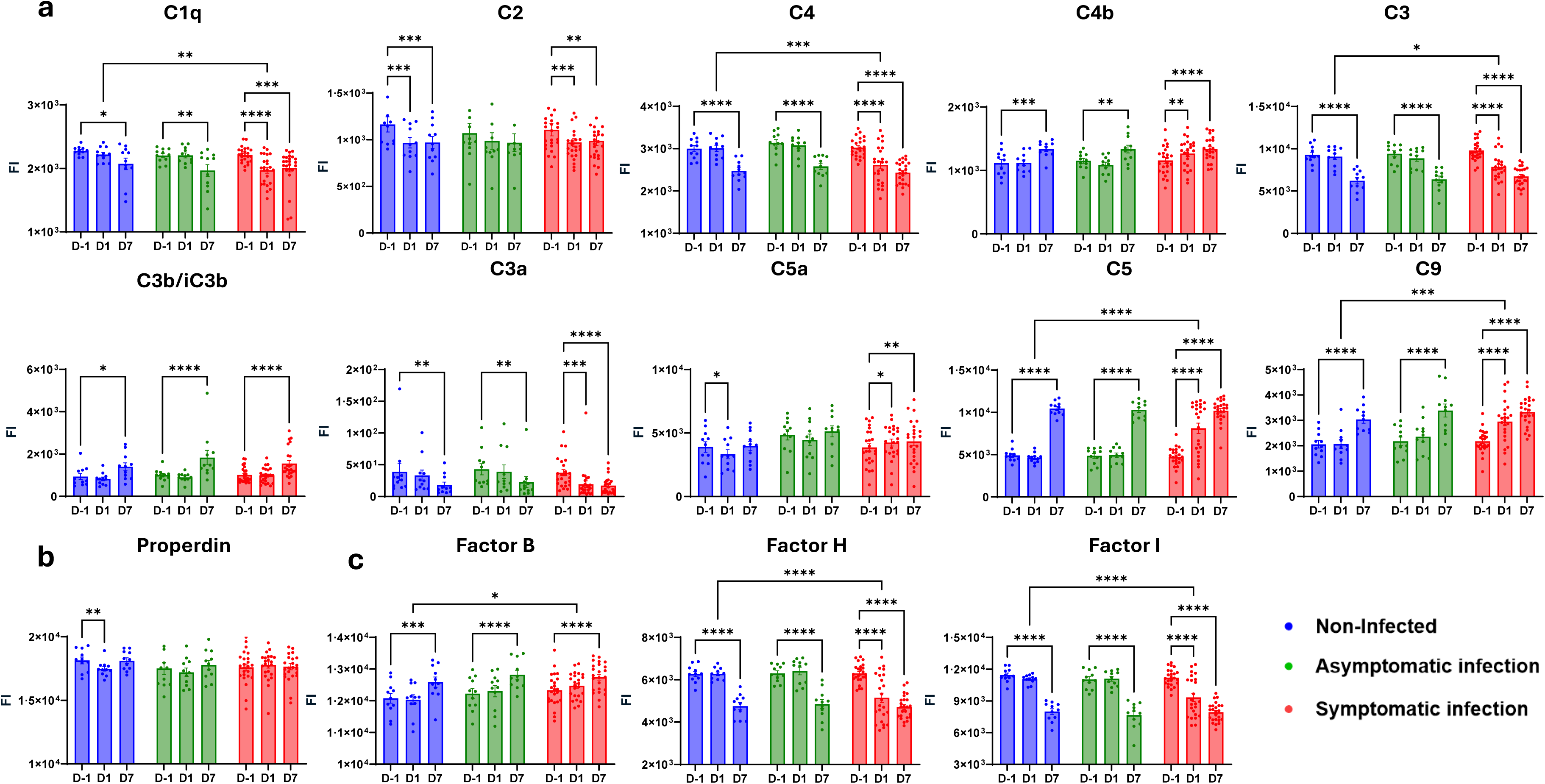
*B. pertussis* challenge induces rapid and robust complement activation in symptomatic participants. Plasma complement proteins measured by Luminex at Day -1 (before challenge) and Days 1 and 7 post-challenge. (a) Complement proteins representing activation of Classic/Lectin complement pathways. (b) Properdin representing engagement of alternative pathway. (c) Factors B, H an I representing complement regulation. Statistical analysis was performed using two-way ANOVA with Bonferroni post test for all complement proteins. Statistical significance is indicated as follows: *p < 0.05, **p < 0.01, ***p < 0.001, ****p < 0.0001.

Notably, the complement cascade is tightly regulated by soluble and membrane-bound inhibitors to ensure that activation and amplification occur only in the appropriate biological context and that host tissues are protected from inadvertent MAC-mediated injury^25^. Consistent with the dynamic of complement activation, consumption of the key inhibitory proteins Factor H and Factor I was evident in symptomatic participants as early as day 1 post-challenge, whereas comparable reductions were delayed until day 7 in non-infected and asymptomatic participants (**Fig.2c**). These findings further highlight marked differences in the timing and magnitude of complement activation among CHIM participants with divergent clinical outcomes. Intriguingly, the magnitude of complement activation was inversely associated with protection, with symptomatic participants exhibiting the most pronounced and early complement responses. These observations suggest that robust complement activation is unlikely to be the primary mechanism underlying spontaneous bacterial clearance and may instead reflect a consequence of uncontrolled infection or heightened inflammation.

### *B. pertussis* induces distinct early mucosal innate immune profiles in CHIM participants

We next measured soluble mediators, including cytokines, chemokines, and cytotoxic effector molecules with antimicrobial activity in NW samples collected at baseline and on days 1 and 3 following *B. pertussis* challenge (**S-Fig.2**). Remarkably, distinct immunological profiles emerged rapidly across clinical outcome groups. Non-infected participants exhibited a significant increase in granzyme A and granzyme B levels in NW at day 3 post-challenge compared to asymptomatic and symptomatic counterparts (**Fig.3a**), providing compelling *in vivo* evidence of *B. pertussis*-induced cytotoxic effector activity in these individuals. In parallel, MCP-2 (CCL8) and IL-29 (IFNL1) were also selectively induced in non-infected individuals at day 3 post-challenge, highlighting an immune profile reminiscent of an antiviral response (**Fig.3a**). In contrast, symptomatic individuals demonstrated rapid upregulation of eotaxin-2 and MIP-1δ (CCL15) at day 1 post challenge, coupled with an increased baseline of ENA-78 (CXCL5) and lymphotactin (XCL1) compared to non-infected and asymptomatic groups (**Fig.3b-3c**). By comparison, asymptomatic participants showed elevated expression of I-TAC (CXCL11), TRAIL (TNFSF10), and CCL28 at day 3 post-challenge (**Fig.3d**). STRING network analysis of these *B. pertussis*-induced soluble mediators further delineated outcome-specific mucosal immune programs (**S-Fig.3**): early cytotoxic signatures predominated in non-infected participants; cytokine/chemokine-mediated inflammatory network characterized symptomatic individuals; and antimicrobial peptide–linked effector pathways were more prominent in asymptomatic participants. Collectively, these data demonstrate that *B. pertussis* induces distinct early mucosal innate immune responses in CHIM participants, with an early cytotoxic signature and an antiviral-like immune profile strongly predicting spontaneous clearance.

**Figure 3:**
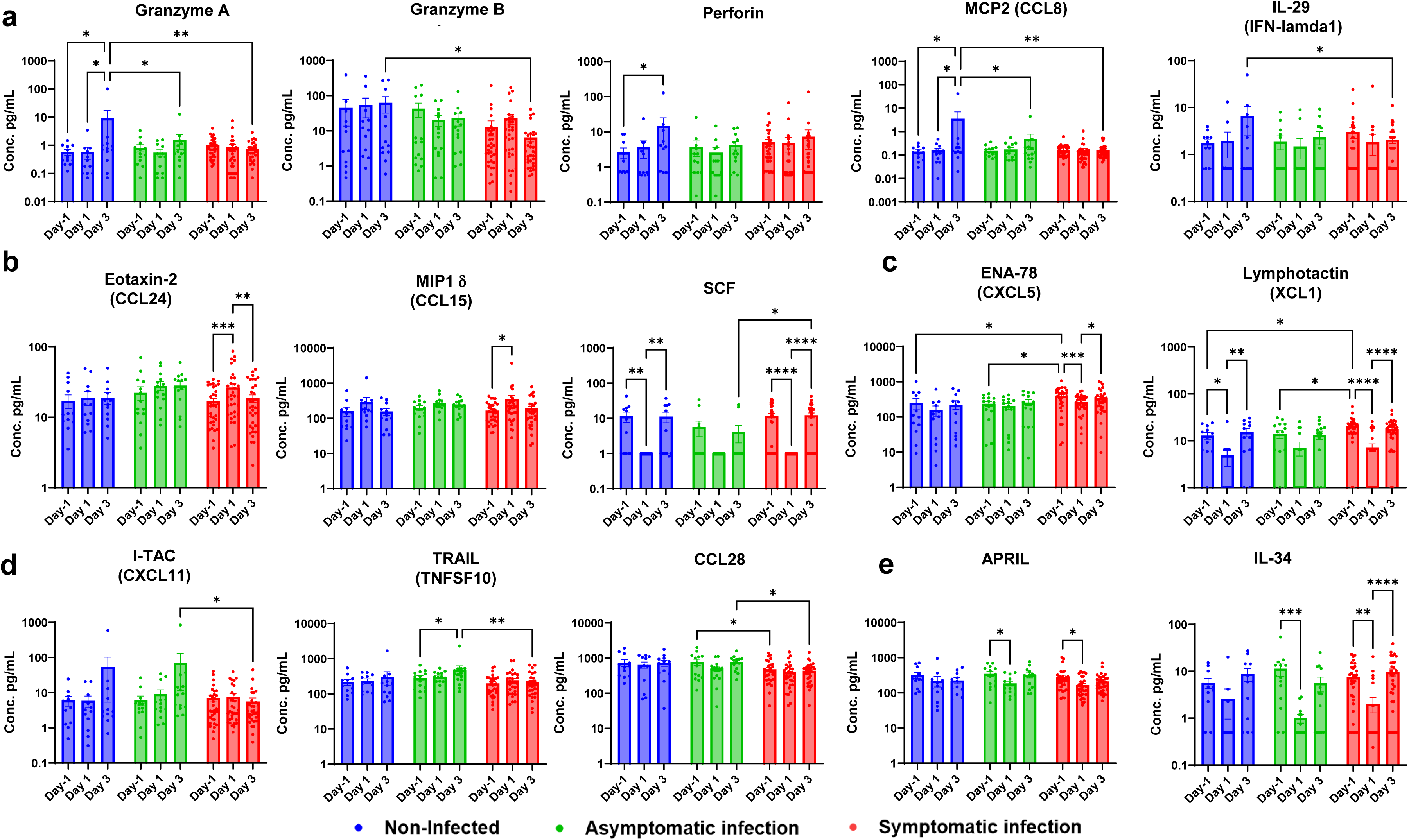
*B. pertussis* elicits distinct mucosal immune responses by clinical outcomes. Levels of soluble mediators were measured by a 48-plex Luminex assay in nasal wash samples collected at Day -1 and Days 1 and 3 post challenge. Data are grouped based on their expression pattern. (a/e) unique for non-infected group, (b/c) unique for symptomatic group, (d) unique for asymptomatic group. Statistical analysis was performed using Repeated two-way ANOVA with Holm-Sidak’s multiple comparisons test for all panels. Statistical significance is indicated as follows: *p < 0.05, **p < 0.01, ***p < 0.001, ****p < 0.0001.

### *B. pertussis* challenge induces characteristic whole-blood innate cellular profiles across three clinical outcome groups

We further characterized *B. pertussis*-induced immune responses by profiling circulating immune cell subsets in fresh whole-blood samples (**Fig.4**) and using paired analyses within individual participants to control for baseline differences across the cohort (**S-Fig.4**). In sharp contrast to the minimal changes observed in B cell and T cell (CD4⁺ and CD8⁺) frequencies and absolute counts (**S-Fig.5a-5c**), characteristic innate immune responses emerged across clinical outcome groups during the first week following *B. pertussis* challenge (**Fig.4**). Notably, non-infected participants exhibited a marked and selective expansion of NK cells, with increases in both frequency and absolute counts at days 3 and 5–7 post-challenge (**Fig.4c**). In contrast, asymptomatic participants showed prominent depletion of circulating neutrophils, likely reflecting their recruitment to the site of infection (**Fig.4a**). Participants who developed symptoms demonstrated broader immune activation, characterized by increased numbers of monocytes (**Fig.4b**) and NK cells (**Fig.4c**), accompanied by a depletion of circulating neutrophils (**Fig.4a**). Elevations in eosinophil frequency and absolute counts were observed in all groups; however, these responses were transient in non-infected and asymptomatic participants, but more pronounced and sustained in symptomatic group (**S-Fig.5d**). Unlike NK cells, the marked expansion of eosinophils was restricted to female participants (**S-Fig.6c)** and was paradoxically observed only in wP immunized individuals (**S-Fig.7c**). MAIT cells showed minimal changes across all groups (**S-Fig.5e**). Taken together with the early cytotoxic signatures observed in non-infected participants (**Fig.3**), these innate cellular profiles support a protective role for NK cells in mediating spontaneous clearance of *B. pertussis*. However, the comparable NK expansion observed in symptomatic participants suggests that the qualitative differences in NK cell responses, rather than numerical expansion alone, may be critical in determining infection outcomes.

**Figure 4:**
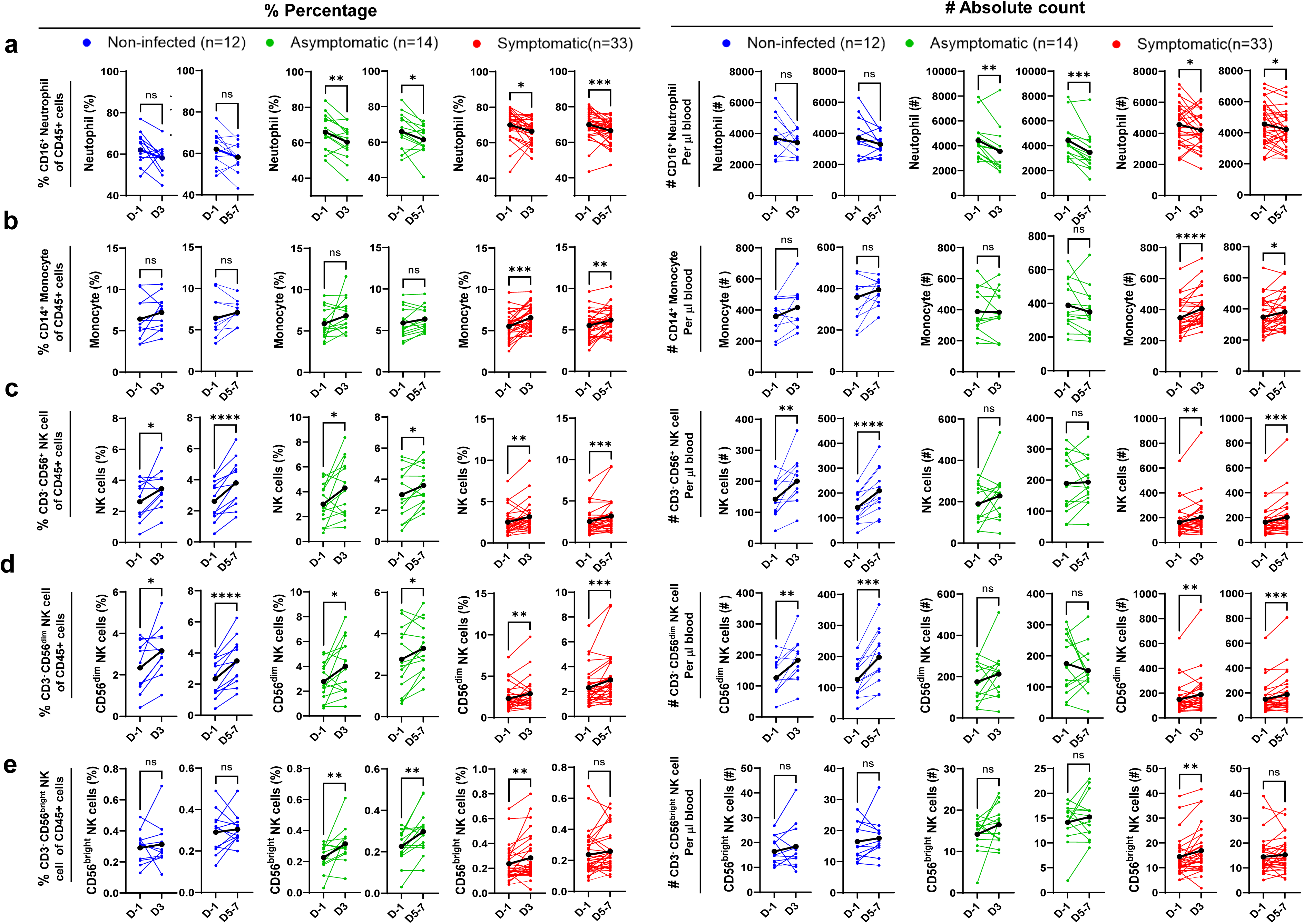
Distinct whole-blood innate immune cellular profiles across three clinical outcome groups following *B. pertussis* challenge. Paired analysis of percentages and absolute counts of different immune subsets at Day −1, Day 3, and Day 5-7 post-challenge. (a) CD45^+^CD16^+^ Neutrophils, (b) CD45^+^CD14^+^ Monocytes, (c) CD45^+^CD3⁻CD56^+^ NK cells, (d) CD56^dim^ NK cells and (e) CD56^bright^ NK cells. A paired t-test or Wilcoxon signed-rank test was used based on the data normality. Statistical significance: *p < 0.05, **p < 0.01, ***p < 0.001, ****p < 0.0001.

Human NK cells are broadly divided into CD56^dim^ and CD56^bright^ subsets, which are conventionally associated with cytotoxicity and cytokine production, respectively.^26^ CD56^bright^ NK cells contain approximately tenfold lower levels of perforin and granzymes than CD56^dim^ cells,^26^ reflecting reduced cytotoxic capacity, but they regulate early immune responses through secretion of IFN-γ and regulatory cytokine IL-10.^27^ Subset analysis revealed selective expansion of the cytotoxic CD56^dim^ NK cells in non-infected participants, whereas symptomatic individuals demonstrated concurrent expansion of both CD56^dim^ and CD56^bright^ populations (**Fig.4d**, **4e**), highlighting that distinct clinical outcomes are likely associated with qualitative differences in NK cells and differential engagement of cytotoxic NK-cell subsets.

### Spontaneous clearance is closely associated with functional maturation of NK cells and the development of adaptive/memory-like NK cells

To further assess qualitative differences of NK-cell responses among different groups, we evaluated *B. pertussis*-induced NK responses *ex vivo* in CHIM participants with distinct clinical outcomes. PBMCs collected 4–6 hours after *B. pertussis* challenge were re-stimulated *in vitro* with HK *B. pertussis*, and the production of the effector molecules including granzyme B, perforin, and IFN-γ by NK cells was assessed by intracellular staining and flow cytometry. Remarkably, t-SNE analysis of NK cells revealed multiple phenotypically distinct subsets. We therefore annotated these NK-cell subsets according to the recently established nomenclature of the human NK-cell atlas (**Fig.5a**, **5b, S-Fig.8**), which was derived from single-cell transcriptomic analyses of human peripheral blood NK cells.^23^ In this framework, six NK subsets (NK2, NKint, NK1A, NK1B, NK1C, and NK3) represent distinct stages of NK-cell maturation and functional specialization. NK2 corresponds to an immature CD56^bright^ regulatory population, NKint represents a transitional state, NK1A/NK1B comprise activated effector subsets, NK1C reflects a highly cytotoxic stage, and NK3 denotes a memory-like NK population associated with prior antigen exposure.^23^ All six subsets were identified in PBMCs both before and after HK *B. pertussis* stimulation and displayed expected patterns of CD16 expression (**Fig.5b**, **5c**). Notably, *in vitro* stimulation with HK *B. pertussis* increased the emerging of NK1A/NK1B subsets compared to unstimulated samples, particularly in non-infected and asymptomatic groups (**Fig.5b**). Contingency analysis of cellular event counts within each subset gate revealed a significant association between clinical outcome and NK-cell subset composition in unstimulated samples, indicating that distinct NK-cell subset distributions were already present among non-infected, asymptomatic, and symptomatic participants at the time of *B. pertussis* challenge (**Fig.5d**). Specifically, NK3 cells were progressively enriched in non-infected and asymptomatic individuals and were least frequent in symptomatic participants (31.1%, 28.4%, and 18.9%, respectively). A similar trend was observed for NK1C subset (26.5%, 18.9%, and 10.8%, respectively). In contrast, NK2 subset was relatively enriched in symptomatic individuals (16.2%) compared with non-infected and asymptomatic groups (both 10.8%). Overall, these data indicate that individuals with non-infected or asymptomatic outcomes are associated with a higher proportion of mature cytotoxic and memory-like NK-cell subsets, whereas symptomatic infection is associated with a relative shift toward less differentiated NK states.

**Figure 5:**
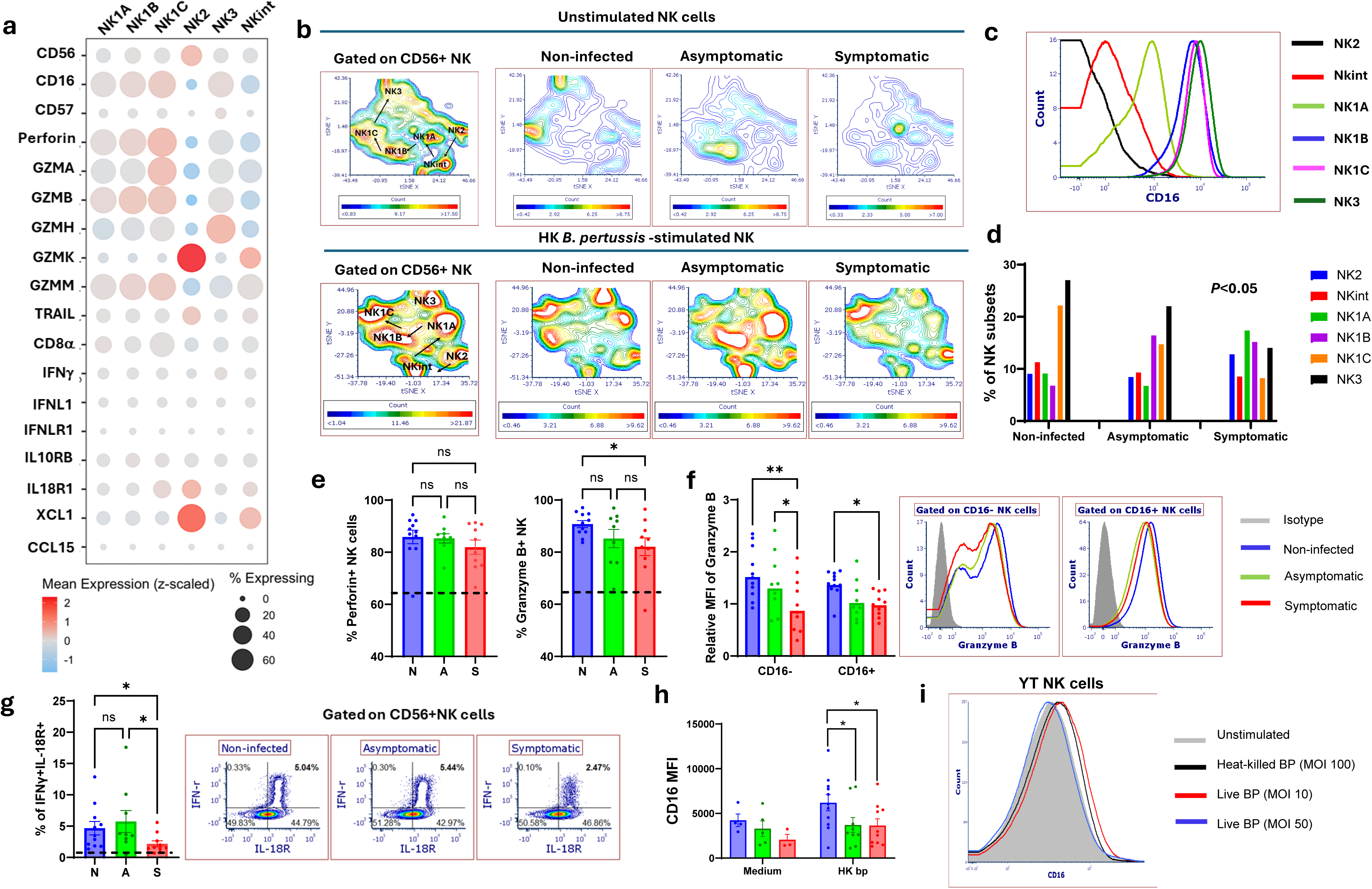
Spontaneous clearance is closely associated with functional maturation of NK cells and the development of adaptive/memory-like NK cells. (a) List of genes expressed by different NK cell subsets obtained from the Human Cell Atlas. (b) t-SNE analysis of unstimulated and HK *B. pertussis*-stimulated CD56⁺ NK cells in PBMC samples. (c) CD16 expression across different NK cell subsets in unstimulated PBMCs. (d) Frequency of different NK cell subsets in unstimulated PBMCs. (e) Percentage of perforin⁺ and granzyme B⁺ CD56⁺ NK cells in HK *B. pertussis*-stimulated PBMC samples with different clinical outcomes. (f) Granzyme B expression (MFI) and representative histograms in CD16⁺ and CD16⁻ NK cells from different clinical outcome groups. Data were pooled from three independent experiments and normalized to the mean expression level of the symptomatic group in each experiment. (g) Frequency and representative plots of IFNγ⁺IL-18R⁺ NK cells in HK *B. pertussis*-stimulated PBMC samples with different clinical outcomes. (h) CD16 MFI in CD56⁺ NK cells with or without HK *B. pertussis* stimulation. (i) CD16 expression in the YT NK cell line following stimulation with either live or heat-killed *B. pertussis*. Dashed lines in panels (e) and (g) indicate expression levels in unstimulated cells. Statistical analyses were performed using Fisher’s exact test for panel (d) using # of events for each gate, Kruskal–Wallis test for panels (e) and (g), and two-way ANOVA with uncorrected Fisher’s LSD post hoc test for panels (f) and (h). Statistical significance is indicated as follows: p < 0.05, p < 0.01, *p < 0.001, and **p < 0.0001; ns: not significant. In panels (e) and (g), N: Non-infected; A: Asymptomatic infection; S: Symptomatic infection.

Notably, NK cells from the non-infected group showed the highest granzyme B production, as assessed by both frequency and median fluorescence intensity (MFI), compared with the symptomatic group (**Fig.5e**, **5f**). These findings were also in strong agreement with the NW Luminex data (**Fig.3**) and aligned perfectly with the NK subset distribution patterns (**Fig.5a-5d**). In addition, HK *B. pertussis* stimulation induced IFN-γ production in NK cells co-expressing IL-18R, a feature enriched in the NK2 subset (**Fig.5a**). Of interest, the frequency of IFN-γ–producing NK cells from the non-infected group was significantly higher than that in symptomatic participants (**Fig.5g**), despite a relative enrichment of NK2 cells in the symptomatic group (**Fig.5d**). These findings suggest that NK2-associated functional responsiveness, in addition to its abundance, also differ between non-infected and symptomatic individuals.

Furthermore, HK *B. pertussis* stimulation increased CD16 expression on cytotoxic CD56^dim^CD16^+^ NK cells in non-infected participants, reaching statistical significance compared with both asymptomatic and symptomatic groups (**Fig.5h**). Given the established role of CD16 in mediating ADCC response, these findings suggest that activated cytolytic NK cells in non-infected individuals possess enhanced potential for ADCC. Of interest, similar CD16 upregulation was also observed in the YT NK cell line following stimulation with either low-dose live bacteria or HK *B. pertussis* **(Fig.5i),** highlighting an intrinsic capacity of cytotoxic NK cells to respond directly to *B. pertussis* stimulation.

### *B. pertussis* directly induces NK cell activation and degranulation

A growing body of evidence demonstrates that NK cells can kill extracellular bacteria and fungi directly or indirectly through interactions with other immune cells^28, 29, 30^. In conjunction with the *B. pertussis*-induced upregulation of CD16 in YT NK cells, the rapid bacterial clearance and pronounced expansion of cytotoxic NK cells observed in non-infected participants suggest that *B. pertussis* may directly initiate NK-cell activation and degranulation. To investigate this mechanism, we assessed degranulation and effector molecule production in the YT NK cell line following *in vitro* stimulation with *B. pertussis*. Remarkably, both live and HK *B. pertussis* induced robust NK cell activation and degranulation, as demonstrated by increased CD107a surface expression (a marker of degranulation) and elevated intracellular levels of granzyme B, perforin, and IFN-γ compared with unstimulated controls (**Fig.6a**). In agreement with these results, YT NK cells primed with HK *B. pertussis* displayed robust inhibitory activity against bacterial growth after 6 hours of co-culture, whereas unprimed YT cells exhibited only modest inhibitory activity (**Fig.6b**).

**Figure 6:**
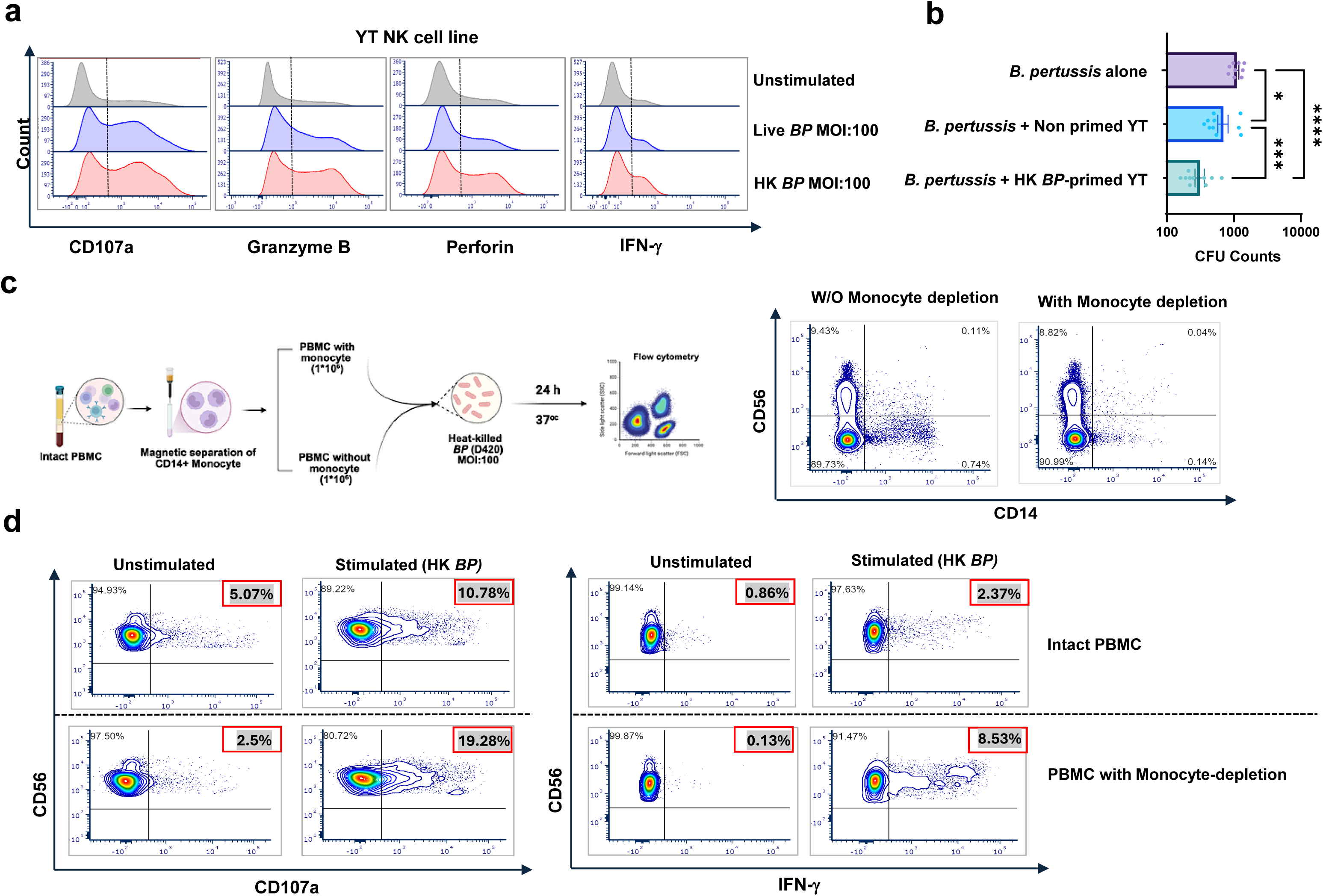
*B. pertussis* directly induces NK cell activation and cytotoxicity. (a) YT NK cell line was stimulated with either live or heat-killed *B. pertussis* (MOI: 100). NK activation and degranulation were assessed by measuring surface CD107a expression and intracellular levels of granzyme B, perforin, and IFN-γ. (b) YT NK cells were primed with heat-killed *B. pertussis at* 100 MOI and subsequently co-cultured with Live *B. pertussis*. Unprimed YT cells and bacteria-only were used as controls. Data were pooled from three independent experiments. (c) Schematic overview of the experimental workflow and confirmation of monocyte depletion from PBMCs. (d) Representative flow cytometry plots showing PBMC with monocytes (top) and PBMC with monocyte-depletion (bottom) under unstimulated and heat-killed *B. pertussis*-stimulated (MOI:100) conditions, demonstrating CD107a degranulation marker and IFN-γ expression in NK cells. Data are representative of two independent experiments.

A previous study demonstrated a critical role of monocytes and macrophages in activating and promoting IFNγ secretion by human NK in the context of *B. pertussis* infection *in vitro.*^31^ However, monocytes may also be hijacked by *B. pertussis* and acquire immunosuppressive properties.^32, 33^ To determine whether NK cell degranulation and cytotoxicity are induced or modulated indirectly through crosstalk with monocytes, we compared NK cell responses in PBMC samples with and without monocyte-depletion (**Fig.6c**). Notably, even incomplete depletion of monocytes augmented NK cell responses to HK *B. pertussis* stimulation, including enhanced CD107a surface expression and increased IFN-γ production relative to intact PBMCs (**Fig.6c**, **6d**). Collectively, these data support a model in which *B. pertussis* directly triggers NK cell activation and degranulation, providing a mechanistic framework for spontaneous clearance observed in non-infected participants.

## DISCUSSION

In this study, we characterized the innate immune landscape of the first North American symptomatic CHIM for *B. pertussis*^22^. In contrast to the asymptomatic CHIM developed by the European PERISCOPE consortium^34^, this model generates a heterogeneous cohort with divergent clinical outcomes, enabling dissection of immune mechanisms underlying sterilizing protection and disease development. Despite the traditionally recognized prolonged incubation period of whooping cough, our analyses revealed that innate immune signatures emerging within 1–3 days after challenge imprint the subsequent trajectory of infection. Symptomatic infection was associated with a type 2 local mucosal cytokine milieu, robust activation of the classical and lectin complement pathways, and broad activation of multiple innate immune cell populations. In contrast, non-infected participants exhibited rapid cytotoxic and antiviral-like effector programming, early engagement of the lectin and alternative complement pathways, and selective activation of NK cells. Importantly, we show that *B. pertussis* directly induces NK-cell activation and degranulation, promoting the production of granzyme B, perforin and IFN-γ together with increased CD16 expression. We further show that NK-cell states follow a maturation hierarchy linking to clinical outcomes, with enrichment of adaptive/memory-like NK cells and highly cytotoxic NK1C subsets in non-infected participants. Collectively, these findings identify NK cell-mediated immunity as an early determinant of resistance to *B. pertussis* infection and support the inclusion of NK-cell functional responses as potential correlates of vaccine-induced protection.

A striking feature distinguishing protected individuals from those who developed infection was the early induction of cytotoxic immune programming in the nasal mucosa and the selective expansion of CD56^dim^CD16^+^ cytotoxic NK cells in peripheral blood within 3 days following challenge (**Fig.3 & Fig.4**). While these findings suggest that rapid activation of cytotoxic NK cells represents a key protective mechanism, it is intriguing why NK cells that expanded in symptomatic participants failed to confer protection. To address this question, we mapped NK-cell subsets according to a recently established NK-cell atlas derived from single-cell transcriptomic analyses of human peripheral blood NK cells.^23^ By integrating seven publicly available datasets, Rébuffet et al. identified three major NK-cell populations, designated NK1, NK2 and NK3, corresponding to conventional CD56^dim^, CD56^bright^ and antigen-experienced adaptive memory-like NK cells, respectively. The NK1 population could be further subdivided into NK1A, NK1B and NK1C subsets, and intermediate NKint population exhibits features of both NK1 and NK2 subsets. Although NK2 displays a distinct ontology from NK1 and NK3, a simplified NK-cell maturation trajectory progresses from NK2 to NKint, then to NK1A/NK1B, followed by NK1C and ultimately NK3. This maturation process is accompanied by progressive downregulation of CD56 and increased expression of CD16 and cytotoxic effector molecules associated with the lytic machinery.^23^ Remarkably, we found that non-infected participants were characterized by enrichment of highly cytotoxic NK1C cells and adaptive memory-like NK3 cells, whereas NK-cell maturation was progressively impaired in asymptomatic participants and further attenuated in symptomatic individuals (**Fig.5b**). *In vitro* analyses further revealed that NK cells from non-infected participants were functionally distinct from those of symptomatic individuals, exhibiting a markedly enhanced responsiveness to *B. pertussis* stimulation, characterized by increased production of NK effector molecules and upregulation of CD16 (**Fig.5e-5h**). In contrast, symptomatic participants were enriched for the NK2 subset, which predominantly expresses granzyme K (**Fig.5a**), a tryptase with known proinflammatory properties but relatively limited cytotoxic activity,^35^ and XCL1 (**Fig.5a**), a chemokine involved in the recruitment of dendritic cells and the promotion of antigen presentation.^36^ Consistent with this observation, baseline XCL1 levels in NW samples from symptomatic participants were significantly higher than those in non-infected and asymptomatic individuals (**Fig.3c**), potentially reflecting the increased abundance of NK2 cells in this group. Currently, the factors underlying the divergent patterns of NK-cell maturation in different clinical groups remain largely unknown. It is also unclear how *B. pertussis* directly activates NK cells and induces degranulation *in vitro*, or whether these responses reflect antigen-specific recognition. Nevertheless, we found that the intrinsic functional differences and maturation states of NK cells in different clinical outcome groups were not attributable to the polymorphisms in killer immunoglobulin-like receptor (KIR) genes (**S-Fig.9**), despite previous studies implicating KIR polymorphisms in host susceptibility to infections.^37, 38^ However, given that 11 of 12 non-infected participants had received wP vaccines, our findings raise the possibility that wP immunization, in contrast to aP vaccines, may preferentially promote NK maturation and the development of adaptive memory-like NK states. Alternatively, the enrichment of mature NK-cell subsets among non-infected participants may reflect broader immunological differences linked to vaccination history rather than a direct effect of wP vaccination per se, and may additionally be shaped by environmental microbial exposures and host microbiota composition. Consistent with this, baseline ENA-78 (CXCL5) levels were elevated in symptomatic participants relative to non-infected and asymptomatic groups (**Fig.3c**). Further studies are warranted to determine how different pertussis vaccine formulations or microbiota species may influence NK-cell maturation and function and whether these effects contribute to long-term protection against *B. pertussis* infection.

In parallel with the induction of cytotoxic mediators, *B. pertussis* challenge also triggered mucosal production of IL-29, a member of the type III interferon (IFN-λ) family known to mediate antiviral and antitumor immunity and to modulate both innate and adaptive immune responses.^39^ Although the precise role of IL-29 in anti-*B. pertussis* immunity remains to be fully elucidated, its selective induction in non-infected participants—but not in asymptomatic or symptomatic individuals—highlights a potentially novel protective role in host defense against bacterial pathogens. Despite the absence of IFN-λ receptor chain 1 (IFN-λR1) expression across NK-cell subsets, IL-29 (IFNL1) transcripts were detected in a fraction of NK cells, including NK1 and NK2 but not NK3, with progressively increased expression across NK1 subsets, reaching the highest levels in the NK1C subset (**S-Fig.10**).^23^ Therefore, the elevated IL-29 levels measured in NW samples from non-infected participants at day 3 post-challenge are likely derived, at least in part, from the mature cytotoxic NK1C subset (**Fig.3a**). Notably, human IL-29 has been shown to suppress Th2 responses, particularly IL-13 secretion, while promoting Th1 immunity.^40, 41^ Similarly, IL-29 has been demonstrated to stimulate IL-12 production in macrophages thereby promoting IFN-γ production by NK cells.^42^ In sharp contrast, eotaxin-2, a potent eosinophil chemoattractant typically induced by Th2 cytokines (IL-4 and IL-13),^43, 44^ was selectively elevated in symptomatic participants shortly after challenge, suggesting a Th2-biased cytokine milieu that favors NK2 differentiation while antagonizing NK1 function. Consistent with this model, NK cells in symptomatic participants exhibited markedly reduced IFN-γ production following *B. pertussis* stimulation. Furthermore, symptomatic participants displayed the most pronounced eosinophilic response in peripheral blood. Collectively, these findings indicate that non-infected and symptomatic individuals are differentially poised to mount Th1/NK1 versus Th2/NK2 inflammatory responses following *B. pertussis* challenge, respectively.

More than 90% of non-infected participants (11/12) were primed with the wP vaccine, whereas nearly 70% of aP-immunized individuals (16/23) developed symptoms, suggesting that prior immunization history likely contributes to the differential Th1/NK1 versus Th2/NK2 responses observed among CHIM participants with distinct clinical outcomes. While it is well established that wP and aP vaccines preferentially induce Th1/Th17 versus Th2 responses, our study provides the first evidence that these formulations may also exert long-lasting effects on NK cell maturation and functionality. Notably, repeated aP vaccination has been reported to modulate host responses to natural *B. pertussis* infection in both animal models^45^ and humans.^46^ Studies in the baboon model indicate that multiple *B. pertussis* exposures are required to retrain immune responses initially primed by aP vaccination^45^, underscoring the durable immunological imprint of prior aP immunization despite declining antibody titers. Therefore, wP and aP vaccines may not only elicit distinct adaptive immune responses but also differentially influence the development or training of memory-like NK cells. In this context, repeated aP immunization may be associated with impaired maturation of cytotoxic NK1 cells and memory-like NK cells, a possibility that warrants further investigation.

In contrast to the protective NK cell-mediated cytotoxic responses, complement activation displayed a paradoxical pattern associated with worse clinical outcomes. Although complement activation became detectable in all participants by day 7 post-challenge, early and pronounced activation of the classical and lectin pathways was observed exclusively in symptomatic participants at day 1 post-challenge. By comparison, non-infected individuals preferentially engaged the lectin and alternative pathways, whereas asymptomatic individuals exhibited minimal complement activation at this early timepoint. These findings argue against a direct role for complement activation in mediating sterilizing protection and instead raise an intriguing question: why was classical complement activation initiated earliest and most strongly in symptomatic participants, despite the significantly higher baseline anti-PT antibody levels observed in non-infected individuals? In addition to antigen-antibody complexes, C-reactive protein (CRP) is known to activate the classical complement pathway through binding to microbial polysaccharides and ligands exposed on damaged cells.^47^ However, baseline CRP levels were comparable across all participants (**S-Fig.11**), making this explanation unlikely. Very recently, Donado and colleagues reported that granzyme K can activate the complement cascade through cleavage of C2 and C4 proteins.^48^. Therefore, one possible explanation for the robust complement activation observed in symptomatic participants is the enrichment of NK2 cells and their increased production of granzyme K upon *B. pertussis* challenge. Under this model, *B. pertussis* challenge directly activates NK cells and induces divergent innate immune programs in different hosts, favoring expansion of mature cytotoxic NK1 subsets in non-infected individuals and NK2 cells in symptomatic participants. Consequently, bacterial burdens diverge rapidly after challenge, leading to greater complement engagement in individuals who develop symptomatic infection through a putative NK2–granzyme K–complement axis. In this context, complement activation may primarily reflect infection progression and tissue damage rather than act as an effector mechanism mediating spontaneous bacterial clearance.

In conclusion, our *in vitro* and *in vivo* findings collectively support a mechanistic model in which divergent early innate immune signatures in both mucosal and systemic compartments are associated with distinct clinical outcomes following *B. pertussis* challenge in CHIM participants (**Graphic abstract**). Most remarkably, the maturation status and subset composition of NK cells appear to play a central role in determining host susceptibility to *B. pertussis* infection. Our findings provide a rationale for the development of NK cell-targeted next-generation pertussis vaccines and underscore the importance of evaluating NK-cell functionality as a potential vaccine-induced correlate of protection.

### Limitations

A limitation of this study is the lack of immune phenotyping of cellular components in NW samples, which precluded direct assessment of innate immune cell dynamics at the mucosal site of infection. Consequently, the cellular sources of soluble mediators and their roles in shaping the mucosal tissue microenvironment remain undefined. Future studies incorporating single-cell transcriptomic profiling of mucosal cellular components will be essential to delineate the host responses that promote sterilizing immunity versus those that drive immunopathology.

## Data Availability

All data produced in the present study are available upon reasonable request to the authors

## ACKNOWLEDGEMENTS

The authors sincerely thank all study participants for their time and commitment. We also acknowledge the contributions of the entire CHIM research team at the Canadian Center for Vaccinology and the Flow Cytometry Core Facility at Dalhousie University for their assistance. The CHIM study was supported by CDC and NIH grants, whereas the work described in this manuscript was supported by the Canadian Institutes of Health Research (CIHR; grant PJT-159700 to JW) and the IWK Health Centre (grant PJ-1029189 to JW). SJ was supported by an IWK Graduate Studentship and a Nova Scotia Graduate Scholarship. HB was supported by a Dalhousie Microbiology and Immunology Graduate Studentship and a Nova Scotia Graduate Scholarship.

## AUTHOR CONTRIBUTIONS

**SJ**: Data acquisition, Data analysis, Visualization, Writing - Original Draft. **ME**: CHIM Methodology, **KLR**: CHIM Methodology. **HB**: Data acquisition, Visualization. **NX**: Methodology, Data acquisition. **EH**: Data acquisition. **CHM**: Methodology. **SH**: CHIM Methodology, Supervision. **JW**: Study Design, Conceptualization, Data analysis, Visualization, Funding Acquisition, Supervision, Writing - Original Draft & Revision. All authors contributed towards the review and editing of the manuscript and agreed with the decision to submit for publication.

## DECLARATION OF INTERESTS

We declare no competing interests.

## Notes

### Competing Interest Statement

The authors have declared no competing interest.

### Clinical Trial

NCT05136599

### Author Declarations

The protocol was approved by the IWK Health Research Ethics Board with written informed consent obtained from all participants

## REFERENCES

1. Kilgore, P.E., Salim, A.M., Zervos, M.J. & Schmitt, H.J. Pertussis: Microbiology, Disease, Treatment, and Prevention. Clinical microbiology reviews 29, 449–486 (2016).

2. Domenech de Cellès, M. & Rohani, P. Pertussis vaccines, epidemiology and evolution. *Nature reviews*. Microbiology 22, 722–735 (2024).

3. Duda-Madej, A., Łabaz, J., Topola, E., Bazan, H. & Viscardi, S. Pertussis-A Re-Emerging Threat Despite Immunization: An Analysis of Vaccine Effectiveness and Antibiotic Resistance. International journal of molecular sciences 26 (2025).

4. Warfel, J.M., Zimmerman, L.I. & Merkel, T.J. Acellular pertussis vaccines protect against disease but fail to prevent infection and transmission in a nonhuman primate model. Proceedings of the National Academy of Sciences of the United States of America 111, 787–792 (2014).

5. Wilk, M.M. et al. Immunization with whole cell but not acellular pertussis vaccines primes CD4 T(RM) cells that sustain protective immunity against nasal colonization with Bordetella pertussis. Emerging microbes & infections 8, 169–185 (2019).

6. Gao, H., Lau, E.H.Y. & Cowling, B.J. Waning Immunity After Receipt of Pertussis, Diphtheria, Tetanus, and Polio-Related Vaccines: A Systematic Review and Meta-analysis. J Infect Dis 225, 557–566 (2022).

7. Klein, N.P. et al. Waning protection following 5 doses of a 3-component diphtheria, tetanus, and acellular pertussis vaccine. Vaccine 35, 3395–3400 (2017).

8. Klein, N.P., Bartlett, J., Rowhani-Rahbar, A., Fireman, B. & Baxter, R. Waning protection after fifth dose of acellular pertussis vaccine in children. The New England journal of medicine 367, 1012–1019 (2012).

9. Warfel, J.M. & Merkel, T.J. Bordetella pertussis infection induces a mucosal IL-17 response and long-lived Th17 and Th1 immune memory cells in nonhuman primates. Mucosal immunology 6, 787–796 (2013).

10. Warfel, J.M., Zimmerman, L.I. & Merkel, T.J. Comparison of Three Whole-Cell Pertussis Vaccines in the Baboon Model of Pertussis. Clinical and vaccine immunology : CVI 23, 47–54 (2016).

11. Borkner, L., Curham, L.M., Wilk, M.M., Moran, B. & Mills, K.H.G. IL-17 mediates protective immunity against nasal infection with Bordetella pertussis by mobilizing neutrophils, especially Siglec-F(+) neutrophils. Mucosal immunology 14, 1183–1202 (2021).

12. Solans, L. et al. IL-17-dependent SIgA-mediated protection against nasal Bordetella pertussis infection by live attenuated BPZE1 vaccine. Mucosal immunology 11, 1753–1762 (2018).

13. Vidarsson, G., Dekkers, G. & Rispens, T. IgG subclasses and allotypes: from structure to effector functions. Frontiers in immunology 5, 520 (2014).

14. Frischauf, N. et al. Complement activation by IgG subclasses is governed by their ability to oligomerize upon antigen binding. Proceedings of the National Academy of Sciences of the United States of America 121, e2406192121 (2024).

15. Diavatopoulos, D.A. & Edwards, K.M. What Is Wrong with Pertussis Vaccine Immunity? Why Immunological Memory to Pertussis Is Failing. Cold Spring Harbor perspectives in biology 9 (2017).

16. Deen, J.L. et al. Household contact study of Bordetella pertussis infections. Clinical infectious diseases : an official publication of the Infectious Diseases Society of America 21, 1211–1219 (1995).

17. Borkner, L., Sutton, C.E., Udayan, S., Jazayeri, S.D. & Mills, K.H.G. Mechanisms of natural and vaccine-induced immunity to Bordetella pertussis. PLoS pathogens 22, e1014128 (2026).

18. McCarthy, K.N., Hone, S., McLoughlin, R.M. & Mills, K.H.G. IL-17 and IFN-γ-producing Respiratory Tissue-Resident Memory CD4 T Cells Persist for Decades in Adults Immunized as Children With Whole-Cell Pertussis Vaccines. J Infect Dis 230, e518–e523 (2024).

19. C, N.C., et al. Acellular Pertussis Vaccines Induce CD8(+) and CD4(+) Regulatory T Cells That Suppress Protective Tissue-Resident Memory CD4(+) T Cells, in Part via IL-10. European journal of immunology 55, e51630 (2025).

20. Netea, M.G. et al. Trained immunity: A program of innate immune memory in health and disease. Science (New York, N.Y.) 352, aaf1098 (2016).

21. Diks, A.M. et al. Distinct early cellular kinetics in participants protected against colonization upon Bordetella pertussis challenge. The Journal of clinical investigation 133 (2023).

22. ElSherif, M.S. et al. A controlled human infection model for symptomatic pertussis in North America using the pertactin-producing clinical isolate D420. 2026.2006.2009.26355227 (2026).

23. Rebuffet, L. et al. High-dimensional single-cell analysis of human natural killer cell heterogeneity. Nature immunology 25, 1474–1488 (2024).

24. Thiriard, A., Raze, D. & Locht, C. Diversion of complement-mediated killing by Bordetella. Microbes and infection 20, 512–520 (2018).

25. Schmidt, C.Q., Lambris, J.D. & Ricklin, D. Protection of host cells by complement regulators. Immunological reviews 274, 152–171 (2016).

26. Jacobs, R. et al. CD56bright cells differ in their KIR repertoire and cytotoxic features from CD56dim NK cells. European journal of immunology 31, 3121–3127 (2001).

27. Poli, A. et al. CD56bright natural killer (NK) cells: an important NK cell subset. Immunology 126, 458–465 (2009).

28. Feehan, D.D. et al. Natural killer cells kill extracellular Pseudomonas aeruginosa using contact-dependent release of granzymes B and H. PLoS pathogens 18, e1010325 (2022).

29. Mody, C.H. et al. Microbial killing by NK cells. Journal of leukocyte biology 105, 1285–1296 (2019).

30. Schmidt, S., Ullrich, E., Bochennek, K., Zimmermann, S.Y. & Lehrnbecher, T. Role of natural killer cells in antibacterial immunity. Expert review of hematology 9, 1119–1127 (2016).

31. Kroes, M.M. et al. Activation of Human NK Cells by Bordetella pertussis Requires Inflammasome Activation in Macrophages. Frontiers in immunology 10, 2030 (2019).

32. Ahmad, J.N. & Sebo, P. Producing Trojans: hijacking of monocyte differentiation by pathogens. Trends in microbiology 34, 378–389 (2026).

33. Ahmad, J.N. et al. Bordetella adenylate cyclase toxin elicits chromatin remodeling and transcriptional reprogramming that blocks differentiation of monocytes into macrophages. mBio 16, e0013825 (2025).

34. de Graaf, H. et al. Controlled Human Infection With Bordetella pertussis Induces Asymptomatic, Immunizing Colonization. Clinical infectious diseases : an official publication of the Infectious Diseases Society of America 71, 403–411 (2020).

35. Turner, C.T. Pro-inflammatory granzyme K contributes extracellularly to disease. Frontiers in immunology 16, 1620670 (2025).

36. Matsuo, K. et al. A Highly Active Form of XCL1/Lymphotactin Functions as an Effective Adjuvant to Recruit Cross-Presenting Dendritic Cells for Induction of Effector and Memory CD8(+) T Cells. Frontiers in immunology 9, 2775 (2018).

37. Middleton, D. & Gonzelez, F. The extensive polymorphism of KIR genes. Immunology 129, 8–19 (2010).

38. Yawata, M., Yawata, N., Abi-Rached, L. & Parham, P. Variation within the human killer cell immunoglobulin-like receptor (KIR) gene family. Critical reviews in immunology 22, 463–482 (2002).

39. Li, M., Liu, X., Zhou, Y. & Su, S.B. Interferon-lambdas: the modulators of antivirus, antitumor, and immune responses. Journal of leukocyte biology 86, 23–32 (2009).

40. Srinivas, S. et al. Interferon-lambda1 (interleukin-29) preferentially down-regulates interleukin-13 over other T helper type 2 cytokine responses in vitro. Immunology 125, 492–502 (2008).

41. Jordan, W.J. et al. Human interferon lambda-1 (IFN-lambda1/IL-29) modulates the Th1/Th2 response. Genes and immunity 8, 254–261 (2007).

42. de Groen, R.A. et al. IFN-λ-mediated IL-12 production in macrophages induces IFN-γ production in human NK cells. European journal of immunology 45, 250–259 (2015).

43. Watanabe, K., Jose, P.J. & Rankin, S.M. Eotaxin-2 generation is differentially regulated by lipopolysaccharide and IL-4 in monocytes and macrophages. Journal of immunology (Baltimore, Md. : 1950) 168, 1911–1918 (2002).

44. Lezcano-Meza, D., Dávila-Dávila, B., Vega-Miranda, A., Negrete-García, M.C. & Teran, L.M. Interleukin (IL)-4 and to a lesser extent either IL-13 or interferon-gamma regulate the production of eotaxin-2/CCL24 in nasal polyps. Allergy 58, 1011–1017 (2003).

45. Kapil, P., Wang, Y., Zimmerman, L., Gaykema, M. & Merkel, T.J. Repeated Bordetella pertussis Infections Are Required to Reprogram Acellular Pertussis Vaccine-Primed Host Responses in the Baboon Model. J Infect Dis 229, 376–383 (2024).

46. Giammanco, A. et al. Analogous IgG subclass response to pertussis toxin in vaccinated children, healthy or affected by whooping cough. Vaccine 21, 1924–1931 (2003).

47. Mold, C., Gewurz, H. & Du Clos, T.W. Regulation of complement activation by C-reactive protein. Immunopharmacology 42, 23–30 (1999).

48. Donado, C.A. et al. Granzyme K activates the entire complement cascade. Nature 641, 211–221 (2025).

