## Supplementary Material and method for "Early Innate Immune Signatures Imprint Clinical Outcomes of *Bordetella pertussis* Challenge in a Controlled Human Infection Model"

### 1. Flowcytometry panel for whole blood staining:

Fresh EDTA-anticoagulated peripheral blood (100 μL) was immunostained with fluorochrome-conjugated antibodies listed in **Suppl. Material and Method Table 1** for 30 minutes at room temperature. Red blood cells were lysed with BD FACS™ Lysing Solution (cat. no. 349202) for 10 minutes, washed with 1X PBS containing 1% FBS, and subsequently fixed with 2% paraformaldehyde. Instrument calibration was performed using SpheroTM Rainbow Calibration Particles (BD, cat. no. 559123) and Anti-Mouse Ig, κ/Negative Control Compensation Particles (BD, cat. no. 51 90-9001229-91). Samples were acquired on a BD LSRFortessa flow cytometer with 200,000 $CD45^+$ events per sample. Data were analyzed using FCS Express 7 software (DeNovo Software, CA). Leukocyte subpopulations were identified within the $CD45^+$ gate as: neutrophils ($CD16^+SSC^{high}$), monocytes ($CD14^+$), NK cells ($CD3^-CD56^+$), MAIT cells ($CD3^+CD8^+CD161^+TCRV\alpha7.2^+$), $CD4^+$ T cells ($CD3^+CD4^+$), $CD8^+$ T cells ($CD3^+CD8^+$), and B cells ($CD3^-CD19^+$). The percentages of each immune subset within $CD45^+$ leukocytes were determined by flow cytometric analysis, and absolute cell counts were calculated using white blood cell counts obtained from the complete blood count (CBC). Eosinophil data were obtained directly from CBC analysis.

**Supplementary Material and Method Table 1:**

| Specificity | Fluorochrome | Clone | Company | Cat# |
|---|---|---|---|---|
| CD45 | FITC | HI30 | BD | 560976 |
| CD3 | BV650 | UCHT1 | BD | 563852 |
| CD4 | BB700 | SK3 | BD | 566392 |
| CD8 | APC-H7 | SK1 | BD | 560179 |
| CD19 | R718 | SJ25C1 | BD | 566946 |
| CD16 | PE-Cy7 | 3G8 | BD | 557744 |
| CD14 | BV786 | M5E2 | BD | 563698 |
| CD56 | AF647 | B159 | BD | 557711 |
| CD161 | PE | DX12 | BD | 556061 |
| TCRVa7.2 | BV421 | OF-5A12 | BD | 749494 |
| Brilliant Stain Buffer Plus | - | - | BD | 566385 |
| Comp Beads, Anti-mouse Ig, k | - | - | BD | 51 90-9001229 |
| Comp Beads, negative control | - | - | BD | 51-90-9001291 |

**2. Flowcytometry panel for intracellular staining:**

Cryopreserved peripheral blood mononuclear cells (PBMC) collected at Day 0 (4-6 h post challenge) were thawed and rested overnight in complete RPMI medium. Cells were seeded at $1\times10^6$ cells per well in round-bottom 96-well plates and stimulated with heat-killed *B. pertussis* strain D420 at a multiplicity of infection (MOI) of 100. After 20 hours of culture at 37°C in 5% $CO_2$, GolgiPlug™ (BD, cat. no.555028) protein transport inhibitor was added, and cells were cultured for an additional 4 hours. Cells were then stained for surface and intracellular markers and analyzed by flow cytometry. In some experiment, monocytes were depleted from PBMC using the EasySep™ Human CD14 Positive Selection Kit II (cat. no. 17858, STEMCELL Technologies) according to the manufacturer's protocol. PBMC samples from healthy donors were processed to generate both monocyte-depleted and intact PBMC populations for the assay. Detailed antibody panel specifications are provided in **Supp. Material and Method Table 2**.

| Specificity | Fluorochrome | Clone | Company | Cat# |
|---|---|---|---|---|
| CD3 | BV650 | UCHT1 | BD | 563852 |
| CD8 | APC-H7 | SK1 | BD | 560179 |
| CD16 | AF700 | 3G8 | BD | 557820 |
| CD56 | AF647 | B159 | BD | 557711 |
| CD161 | PE | DX12 | BD | 556061 |
| TCRVa7.2 | BV421 | OF-5A12 | BD | 749494 |
| IFN-γ | FITC | B27 | BD | 552887 |
| Perforin | PE-CF594 | δG9 | BD | 563763 |
| Granzyme B | PE-Cy7 | QA16A02 | BD | 372214 |
| CD107a | BB700 | H4A3 | BD | 566556 |
| Horizon™ Fixable Viability Stain 575V | FVS575V | - | BD | 565694 |
| Cytofix/Cytoperm™ Plus | - | - | BD | 555028 |
| Brilliant Stain Buffer Plus | - | - | BD | 566385 |
| eBioscience™ Monensin Solution (1000X) | - | - | eBioscience | 00-4505-51 |
| Comp Beads, Anti-mouse Ig, k | - | - | BD | 51 90-9001229 |
| Comp Beads, negative control | - | - | BD | 51-90-9001291 |

**Luminex multiplex**

Nasal wash samples collected at baseline (Day-1) and on Days 1 and 3 post-challenge were measured using the Human Cytokine Panel B 48-Plex Discovery Assay® (MilliporeSigma, Burlington, Massachusetts, USA) on a Luminex™ 200 system (Luminex, Austin, TX, USA) by Eve Technologies Corp (Calgary, Alberta). The assay sensitivities for the analytes ranged from 0.05-100 pg/mL. For analyte concentrations below the limit of detection, a value corresponding to 50% of the lower detection limit was entered for statistical analysis. Data are reported as analyte concentrations in pg/mL.

Complement components were measured in plasma samples collected at baseline (Day −1) and on Days 1 and 7 post-challenge using the MILLIPLEX® Human Complement Expanded Magnetic Bead Panel 1 (cat. no. HCMPEX11-19K), Panel 2 (cat. no. HCMP2MAG-19K), and HU C3a SIMPLEX assay (cat. no. EPX010-12282-901) according to the manufacturer's instructions. Data are presented as fluorescence intensity (FI) values for each analyte.

**Degranulation assay using YT NK Cell Line**

The YT NK cell line was a kind gift from Dr. Christopher Mody (University of Calgary). The YT NK cell line was maintained in RPMI medium 1640 1x (Gibco, Cat. No. 22400-089) supplemented with 10% fetal bovine serum (FBS), 1% penicillin-streptomycin, 1% sodium pyruvate (Cat. No. 11360-070), and 1% MEM non-essential amino acids (NEAA; Cat. No. 11140-050). In order to test *B. pertussis*-induced NK activation and degranulation, YT NK cells ($1\times10^6$) were stimulated with live or heat-killed *B. pertussis* strain D420 at MOI of 10,50, and 100 in the presence of anti-CD107a antibody for 1 hour at 37°C with 5% $CO_2$. GolgiPlug™ and GolgiStop™ protein transport inhibitors were then added, and cells were incubated for an additional 4 hours. Cells were subsequently stained for surface markers (CD16, CD56) and intracellular molecules (IFN-γ, perforin, granzyme B) using flow cytometry (**Supp Material and Method Table 2**). Experiment was performed in triplicate and repeated independently twice.

**NK Cell-Mediated Bacterial Killing Assay**

To assess the cytotoxic activity of YT NK cells against *B. pertussis*, YT cells cultured in complete RPMI medium were harvested and washed three times with 1× DPBS (cat. no. 14040-117) to remove residual antibiotics. For NK-cell priming, approximately $5 \times 10^6$ YT cells were incubated with heat-killed *B.*

*pertussis* at MOI 100 for 6 h at 37°C in 5% $CO_2$ in 200 μL DPBS. Unprimed YT cells were incubated under identical conditions in the absence of bacteria. Following the 6-h priming phase, live *B. pertussis* was mixed with either primed or unprimed YT cells at various effector-to-bacterium ratios and cultured at 37°C in 5% $CO_2$. After 6 h of co-culture, a 10-fold volume of sterile water was added to lyse YT cells, and appropriate volumes of the resulting suspensions were plated onto charcoal agar plates and incubated at 37°C for 5–7 days. Live *B. pertussis* subjected to the same procedure but cultured in the absence of YT cells served as controls. Bacterial colonies were enumerated and expressed as colony-forming units (CFUs). Each condition was tested in triplicate, and the experiment was independently repeated three times.

**KIR genotyping**

Genomic DNA was isolated from PBMCs samples of 50 participants. KIR genotyping was performed by PCR using AccuStart II GelTrack® PCR SuperMix (Quantabio; cat. no. 95136-500) and gene-specific primer pairs detailed in **Supplementary Material and Method Table 3**. PCR products were analyzed by agarose gel electrophoresis to determine the presence or absence of individual KIR genes. Homozygous and heterozygous gene copy numbers were inferred from the size and relative intensity of the amplified DNA bands as described previously.[1]

1. Vilches, C., Castaño, J., Gómez-Lozano, N. & Estefanía, E. Facilitation of KIR genotyping by a PCR-SSP method that amplifies short DNA fragments. Tissue antigens 70, 415-422 (2007).

**Supp. table 3:** Human KIR genes primer sequence

| Reaction | Genea | Name | Forward primer/ Sequence (5#–3#) | Nameb | Reverse primer/ Sequence (5#–3#) | bp | Name | Forward primer/ Sequence (5#–3#) | bp | Company |
|---|---|---|---|---|---|---|---|---|---|---|
| 1 | KIR2DL1 | Fa517d | gttggtcagatgtcatgtttgaa | Rc621d | cctgccaggtcttgcg | 142 | | | | IDT |
| 2 | KIR2DL2 | Fcon750d | aaaccttctctctcagccca | Rt854 | gccctgcagagaacctaca | 142 | | | | IDT |
| 3 | KIR2DL3 | Fcon1254d | agaccctcaggaggtga | Rt1375 | caggagacaactttggatca | 156 | | | | IDT |
| 4 | KIR3DL1 | Ft624 | ccatyggtcccatgatgct | Rt697d | ccacgatgtccagggga | 108 | Ftt624d | tccatcggtcccatgatgtt | 109 | IDT |
| 5 | KIR3DL2 | Fg864d | catgaacgtaggctccg | Rc962 | gaccacacgcagggcag | 131 | | | | IDT |
| 6 | KIR2DS1 | Fg621 | tctccatcagtcgcatgag | Rcon682d | ggtcactgggagctgac | 96 | Fa621b | tctccatcagtcgcatgaa | 96 | IDT |
| 7 | KIR2DS2 | Fa546 | tgcacagagaggggaagta | Rcon621d | ccctgcaaggtcttgca | 110 | | | | IDT |
| 8 | KIR2DS3 | Ft803d | cttgtcctgcagctcct | Ra925 | gcatctgtaggttcctcct | 158 | | | | IDT |
| 9 | KIR2DS4 | Fat781d | ggttcaggcaggagagaat | Rca877d | ctggaatgttccgtkgatg | 133/111 | | | | IDT |
| 10 | KIR2DS5 | Fc551 | agagaggggacgtttaacc | Rcon662d | ctgatagggggagtgagt | 147 | | | | IDT |
| 11 | KIR3DS1 | Fg624d | catcggttccatgatgcg | Rt697d | ccacgatgtccagggga | 107 | Fg624bd | catcagttccatgatgcg | 107 | IDT |
| 12 | KIR3DL3 | Fg510d | aatgttggtcagatgtcag | Rta669bd | gcygacaactcatagggta | 196 | | | | IDT |
| 13 | KIR3DX1 | Fma920d | tttctgtgggccgtgcaa | Rdel967bd | gtcactgggggcttatag | 88 | | | | IDT |
| 14 | KIR2DL4 | Ftgc157bd | tcaggacaagcccttctgc | Rga250bd | ggacagggaccccatctttc | 131 | | | | IDT |
| 15 | KIR2DL5 | Fag843d | atctatccagggaggggag | Rc953d | catagggtgagtcatggag | 147 | | | | IDT |
| 16 | Internal positive control HLA-DRA gene | FDRA360 | gaggtaactgtgctcacgaacagc | RDRA595 | ggtccataccccagtgcttgagaag | For reactions 1–15 (product length: 283 bp) | RDRA633 | cacgttctctgtagtctctggg | For reaction 16 (product length: 608 bp) | IDT |

**Supp. Table 1:** Demographic characteristics by dose group and sex. Data are n (%) or mean ± SD

| Demographic characteristics | | Total (n=59) | Challenge dose | | | | |
|---|---|---|---|---|---|---|---|
| | | | $10^6$ CFU (n=6) | $5\times10^6$ CFU (n=10) | $10^7$ CFU (n=22) | $5\times10^7$ CFU (n=12) | $10^8$ CFU (n=9) |
| Sex and original pertussis vaccine | **Male, n (%)** | 34 (57.63%) | 4 | 6 | 14 | 5 | 5 |
| | Age, Mean ± SD | 27.68 ± 5.19 | 30.5 ± 3.87 | 28.66 ± 4.22 | 27.57 ± 6.5 | 27.4 ± 4.27 | 24.8 ± 3.27 |
| | wP Vaccine, n (%) | 21 (61.76%) | 3 (75.0%) | 5 (83.3%) | 8 (57.0%) | 4 (80.0%) | 1 (20.0%) |
| | aP Vaccine, n (%) | 13 (38.24%) | 1 (25.0%) | 1 (16.7%) | 6 (43.0%) | 1 (20.0%) | 4 (80.0%) |
| | **Female, n (%)** | 25 (42.37%) | 2 | 4 | 8 | 7 | 4 |
| | Age, Mean ± SD | 28.20 ± 6.51 | 34.5 ± 2.12 | 26.25 ± 4.5 | 24.37 ± 6.09 | 30.14 ± 7.4 | 31.25 ± 5.43 |
| | wP Vaccine, n (%) | 15 (60.0%) | 2 (100%) | 3 (75.0%) | 3 (37.5%) | 4 (57.14%) | 3 (75.0%) |
| | aP Vaccine, n (%) | 10 (40.0%) | 0 (0.0%) | 1 (25.0%) | 5 (62.5%) | 3 (42.86%) | 1 (25.0%) |
| | wP - Age, Mean ± SD | 31.47 ± 4.31 | 33.2 ± 1.92 | 29.25 ± 3.11 | 31.36 ± 5.37 | 32.13 ± 5.17 | 32.75 ± 3.10 |
| | aP - Age, Mean ± SD | 22.30 ± 1.92 | 25 ± 0.0 | 21.5 ± 0.71 | 21.45 ± 2.07 | 22.75 ± 0.96 | 23.60 ± 1.52 |
| Clinical Outcomes (by Sex and original Pertussis vaccine) | **Non-infected** | | | | | | |
| | Male, n (%) | 9 (26.47%) | 2 (50.0%) | 4 (66.6%) | 2 (14.3%) | 1 (20.0%) | 0 (0.0%) |
| | Female, n (%) | 3 (12.0%) | 1 (50.0%) | 0 (0.0%) | 1 (12.5%) | 0 (0.0%) | 1 (25.0%) |
| | wP , n (%) | 11 (30.56 %) | 3 (60.0%) | 4 (50.0%) | 2 (18.18%) | 1 (12.5%) | 1 (25.0%) |
| | aP , n (%) | 1 (4.35%) | 0 (0.0%) | 0 (0.0%) | 1 (9.09%) | 0 (0.0%) | 0 (0.0%) |
| | **Asymptomatic infection** | | | | | | |
| | Male , n (%) | 10 (29.42%) | 0 (0.0%) | 1 (16.7%) | 3 (21.4%) | 3 (60.0%) | 3 (80.0%) |
| | Female , n (%) | 4 (16.0%) | 0 (0.0%) | 3 (75.0%) | 0 (0.0%) | 1 (14.3%) | 0 (0.0%) |
| | wP , n (%) | 8 (22.2%) | 0 (0.0%) | 3 (37.5%) | 2 (18.18%) | 3 (37.5%) | 0 (0.0%) |
| | aP , n (%) | 6 (26.08%) | 0 (0.0%) | 1 (50.0%) | 1 (9.09%) | 1 (50.0%) | 3 (60.0%) |
| | **Symptomatic infection** | | | | | | |
| | Male , n (%) | 15 (44.11%) | 2 (50.0%) | 1 (16.7%) | 9 (64.3%) | 1 (20.0%) | 2 (40.0%) |
| | Female , n (%) | 18 (72.0%) | 1 (50.0%) | 1 (25.0%) | 7 (87.5%) | 6 (85.7%) | 3 (75.0%) |
| | wP , n (%) | 17 (47.22) | 2 (40.0%) | 1 (12.5%) | 7 (63.64%) | 4 (50.0%) | 3 (75.0%) |
| | aP , n (%) | 16 (69.57%) | 1 (100%) | 1 (50.0%) | 9 (81.82%) | 3 (75.0%) | 2 (40.0%) |

**Supp. Table 2:** Rates of infection and clinical symptoms following *B. pertussis* challenge

| **Dose** | **N** | **Total infected** | *Asymptomatic Infection* | *Symptomatic Infection* | **Non-infected** |
|---|---|---|---|---|---|
| $10^4$ | 6 | 0 (0.0%) | *0 (0.0%)* | *0 (0.0%)* | 6 (100.0%) |
| $10^5$ | 5 | 2 (40.0%) | *1 (20.0%)* | *1 (20.0%)* | 3 (60.0%) |
| $5 \times 10^5$ | 5 | 1 (20.0%) | *0 (0.0%)* | *1 (20.0%)* | 4 (80.0%) |
| $10^6$ | 6 | 3 (50.0%) | *0 (0.0%)* | *3 (50.0%)* | 3 (50.0%) |
| $5 \times 10^6$ | 10 | 6 (60.0%) | *4 (40.0%)* | *2 (20.0%)* | 4 (40.0%) |
| $10^7$ | 22 | 19 (86.4%) | *3 (13.6%)* | *16 (72.7%)* | 3 (13.6%) |
| $5 \times 10^7$ | 12 | 11 (91.7%) | *4 (33.3%)* | *7 (58.3%)* | 1 (8.3%) |
| $10^8$ | 9 | 8 (88.9%) | *3 (33.3%)* | *5 (55.6%)* | 1 (11.1%) |

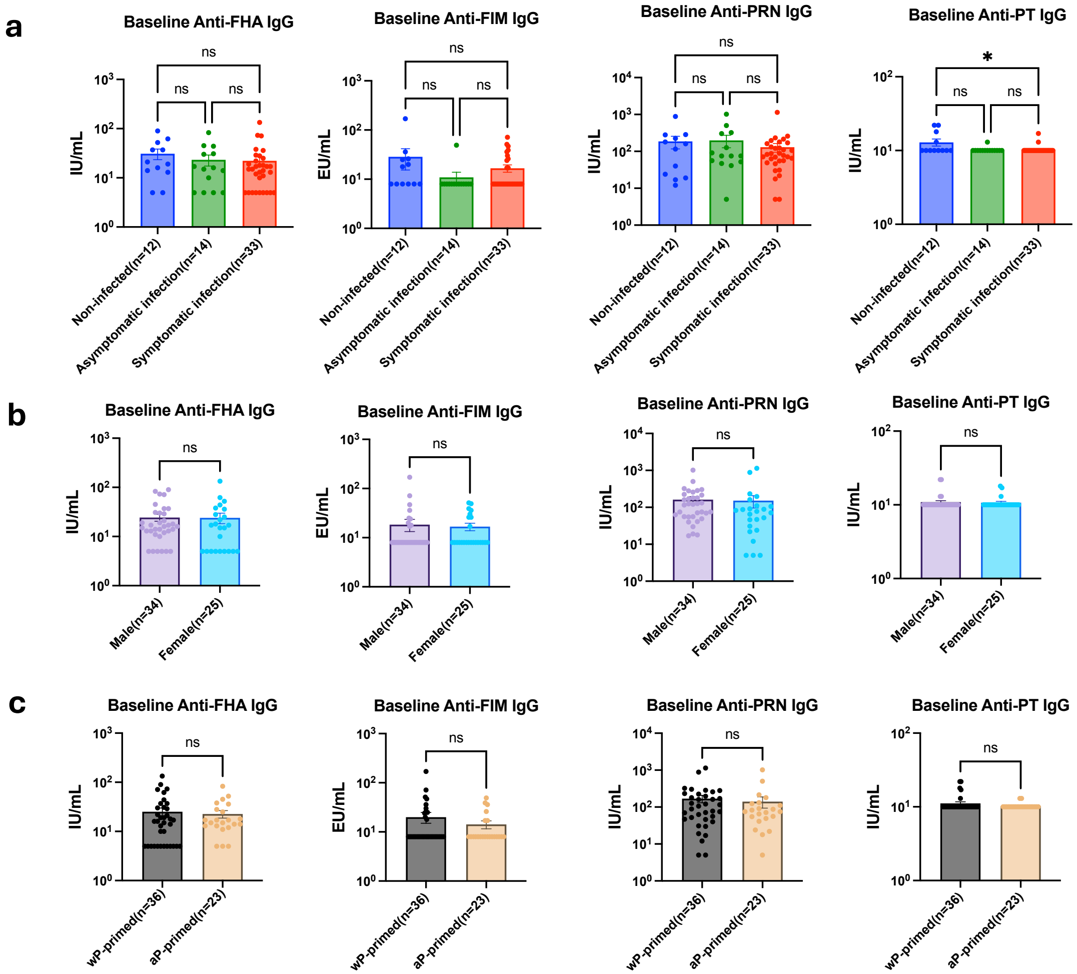


**Supp. Fig. 1: Baseline serum pertussis-specific antibody levels across clinical outcome groups.** Serum antibody concentrations were measured at baseline by ELISA against four *B. pertussis* antigens: pertactin (PRN), filamentous hemagglutinin (FHA), pertussis toxin (PT), and fimbriae (FIM). Comparisons are shown for **(a)** three clinical outcome groups (non-infected, asymptomatic infection, and symptomatic infection), **(b)** biological sex (male vs. female), and **(c)** original vaccination history (whole-cell pertussis vaccine, wP vs. acellular pertussis vaccine, aP). Data are presented as mean ± SEM. Statistical comparisons were performed using one-way ANOVA (Kruskal-Wallis test) or unpaired t-test(Mann-Whitney test) where appropriate. ns, not significant; *p < 0.05.

| pg/ml | Non-infected Day-1 | Non-infected Day 1 | Non-infected Day 3 | Asymptomatic Day-1 | Asymptomatic Day 1 | Asymptomatic Day 3 | Symptomatic Day-1 | Symptomatic Day 1 | Symptomatic Day 3 |
|---|---|---|---|---|---|---|---|---|---|
| 6CKine | 124.4 | 102.1 | 123.4 | 102.7 | 48.0 | 68.1 | 102.0 | 51.9 | 84.4 |
| APRIL | 320.8 | 218.9 | 223.9 | 338.8 | 186.2 | 310.8 | 279.3 | 166.6 | 211.6 |
| BAFF | 4.5 | 3.4 | 9.3 | 5.7 | 2.6 | 5.0 | 8.7 | 3.3 | 11.1 |
| BCA-1 | 5.1 | 4.9 | 8.4 | 17.4 | 9.4 | 12.1 | 7.2 | 10.9 | 7.5 |
| CCL28 | 735.5 | 644.7 | 719.3 | 744.3 | 502.6 | 757.6 | 486.1 | 401.6 | 442.5 |
| CTACK | 1.2 | 1.4 | 1.5 | 1.1 | 0.9 | 0.9 | 1.4 | 1.6 | 1.2 |
| CXCL16 | 11.0 | 10.4 | 3.9 | 5.5 | 1.8 | 1.0 | 3.7 | 2.7 | 1.3 |
| ENA-78 | 249.2 | 156.4 | 225.4 | 254.8 | 218.5 | 258.6 | 391.1 | 259.6 | 348.3 |
| Eotaxin-2 | 17.1 | 19.0 | 18.8 | 22.2 | 27.9 | 27.1 | 16.8 | 26.5 | 18.8 |
| Eotaxin-3 | 1.0 | 0.9 | 0.8 | 0.7 | 0.7 | 1.2 | 1.4 | 0.8 | 0.9 |
| GCP-2 | 62.9 | 47.4 | 56.1 | 75.3 | 88.8 | 83.8 | 93.1 | 82.1 | 111.1 |
| Granzyme A | 0.6 | 0.6 | 9.1 | 0.8 | 0.6 | 1.5 | 1.0 | 0.9 | 0.8 |
| Granzyme B | 45.2 | 54.4 | 62.9 | 40.2 | 19.7 | 21.4 | 13.4 | 22.7 | 6.6 |
| HMGB1 | 309.3 | 30.0 | 661.7 | 378.5 | 71.2 | 333.5 | 543.3 | 412.6 | 648.6 |
| I-309 | 0.1 | 0.1 | 0.3 | 0.1 | 0.1 | 0.1 | 0.4 | 0.3 | 0.2 |
| I-TAC | 6.0 | 5.9 | 54.0 | 6.3 | 8.9 | 66.1 | 7.1 | 7.7 | 5.8 |
| IFNβ | 0.1 | 0.3 | 0.3 | 0.1 | 0.1 | 0.1 | 0.3 | 0.2 | 0.2 |
| IFNω | 0.6 | 0.7 | 0.9 | 0.8 | 0.7 | 0.8 | 1.0 | 0.9 | 0.8 |
| IL-11 | 2.1 | 1.6 | 1.1 | 0.9 | 0.2 | 0.3 | 1.3 | 0.8 | 0.5 |
| IL-16 | 15.1 | 14.2 | 31.0 | 47.2 | 36.0 | 35.7 | 39.5 | 49.1 | 95.3 |
| IL-20 | 3.0 | 3.4 | 5.6 | 2.9 | 1.6 | 2.2 | 5.9 | 5.0 | 3.6 |
| IL-21 | 2.2 | 3.3 | 2.9 | 1.4 | 0.4 | 1.2 | 3.8 | 2.4 | 2.1 |
| IL-23 | 32.6 | 48.3 | 35.3 | 19.4 | 6.0 | 9.9 | 44.1 | 38.6 | 20.2 |
| IL-24 | 8.5 | 9.0 | 14.3 | 6.5 | 1.7 | 4.5 | 20.4 | 8.4 | 12.6 |
| IL-28A | 2.8 | 1.6 | 3.7 | 2.1 | 1.0 | 1.5 | 3.8 | 1.7 | 3.0 |
| IL-29 | 1.7 | 1.9 | 6.5 | 1.8 | 1.4 | 2.2 | 3.1 | 1.9 | 2.1 |
| IL-31 | 1.0 | 1.3 | 1.4 | 1.1 | 0.6 | 1.0 | 1.6 | 1.3 | 2.7 |
| IL-33 | 9.7 | 6.2 | 20.6 | 12.1 | 1.8 | 4.3 | 20.2 | 9.1 | 11.2 |
| IL-34 | 5.7 | 2.6 | 8.8 | 10.7 | 1.0 | 5.9 | 7.7 | 2.1 | 9.5 |
| IL-35 | 8.7 | 10.7 | 11.3 | 8.3 | 6.2 | 7.0 | 12.9 | 12.9 | 13.2 |
| LIF | 1.1 | 1.9 | 2.1 | 1.1 | 0.6 | 1.1 | 2.3 | 1.9 | 1.1 |
| Lymphotactin | 12.9 | 4.9 | 15.2 | 14.4 | 8.0 | 13.6 | 20.7 | 6.8 | 17.9 |
| MCP-2 | 0.1 | 0.2 | 3.6 | 0.2 | 0.2 | 0.5 | 0.2 | 0.2 | 0.2 |
| MCP-4 | 0.9 | 0.5 | 1.0 | 0.6 | 0.3 | 0.5 | 1.1 | 0.6 | 0.8 |
| MIP 1δ | 159.8 | 288.5 | 156.1 | 195.4 | 272.8 | 236.0 | 166.7 | 348.6 | 193.4 |
| MIP-3α | 0.8 | 1.5 | 1.2 | 1.0 | 1.3 | 2.6 | 1.9 | 1.4 | 1.6 |
| MIP-3β | 4.6 | 3.0 | 7.3 | 5.1 | 5.4 | 9.2 | 5.8 | 3.6 | 7.9 |
| MPIF-1 | 1.1 | 1.3 | 1.5 | 1.2 | 1.1 | 1.4 | 1.2 | 1.1 | 1.0 |
| Perforin | 2.6 | 3.6 | 14.6 | 3.8 | 2.4 | 4.0 | 5.1 | 4.9 | 7.4 |
| sCD137 | 0.3 | 0.4 | 0.6 | 0.5 | 0.4 | 0.5 | 0.8 | 0.7 | 0.7 |
| SCF | 11.4 | 1.0 | 11.2 | 5.4 | 1.0 | 5.0 | 12.0 | 1.0 | 12.1 |
| SDF-1 | 44.0 | 29.9 | 46.7 | 47.3 | 39.6 | 51.1 | 54.0 | 40.0 | 52.9 |
| sFas | 0.2 | 0.3 | 0.3 | 0.4 | 0.4 | 0.5 | 0.4 | 0.4 | 0.4 |
| sFasL | 0.7 | 0.7 | 3.2 | 0.5 | 0.1 | 0.8 | 2.5 | 2.3 | 2.4 |
| TARC | 1.8 | 1.3 | 1.9 | 1.5 | 1.5 | 1.9 | 1.6 | 1.7 | 2.0 |
| TPO | 0.3 | 0.7 | 0.5 | 0.2 | 0.2 | 0.2 | 1.2 | 1.5 | 0.5 |
| TRAIL | 214.4 | 230.9 | 298.4 | 278.7 | 299.0 | 454.1 | 196.1 | 244.0 | 214.9 |
| TSLP | 0.2 | 0.1 | 0.3 | 0.2 | 0.1 | 0.2 | 0.3 | 0.2 | 0.2 |

Color scale (pg/ml): 200, 400, 600

**Supp. Fig. 2: cytokines and chemokines in nasal wash samples in 3 different clinical outcome groups.** Heatmap showing the average nasal wash analyte levels measured by the Luminex 48-Plex Discovery Assay at Day −1 (baseline), Day 1, and Day 3 following challenge in each study group. Colors represent mean analyte concentrations (pg/mL).

**STRING analysis in non-infected**

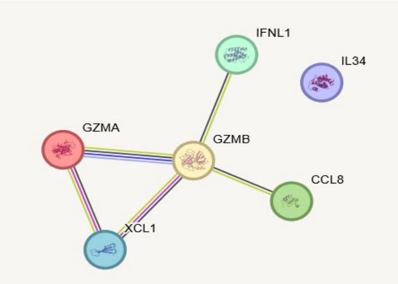


| GO-term | Description | FDR |
|---|---|---|
| GO:0140507 | Granzyme-mediated programmed cell death | 0.0061 |
| GO: 0001906 | Cell killing | 0.0076 |
| GO: 0019221 | Cytokine-mediated signaling pathway | 0.0036 |
| GO: 0002687 | Positive regulation of leukocyte migration | 0.0111 |

**STRING analysis in asymptomatic infection**

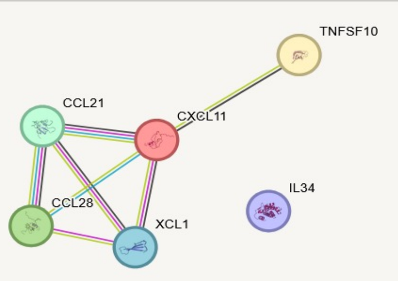


| GO-term | Description | FDR |
|---|---|---|
| GO:0031640 | Killing of cells of another mechanism | 4.53e-5 |
| GO: 0061844 | Antimicrobial peptide-mediated humoral immune response | 0.00014 |
| GO: 0071622 | Regulation of granulocyte chemotaxis | 0.00076 |
| GO: 0048247 | Lymphocyte chemotaxis | 0.00076 |

**STRING analysis in symptomatic infection**

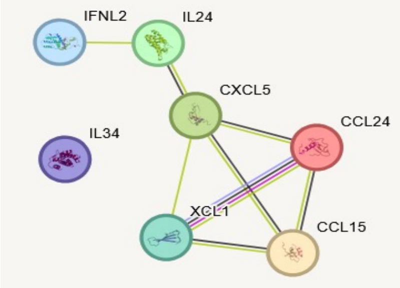


| GO-term | Description | FDR |
|---|---|---|
| GO:0030593 | Neutrophil chemotaxis | 2.53e-5 |
| GO: 0070098 | Chemokine-mediated signaling pathway | 2.53e-5 |
| GO: 0019221 | Cytokine-mediated signaling pathway | 1,64e-6 |
| GO: 0002548 | Monocyte chemotaxis | 0.00031 |

**Supp. Fig. 3: String analysis of mucosal immune responses induced by *B. pertussis* challenge:** Protein–protein interaction networks (left) and Gene Ontology (GO) enrichment analyses (right) of differentially expressed soluble mediators identified in three clinical groups. Molecules include Granzyme A (GZMA), Granzyme B (GZMB), X-C Motif Chemokine Ligand 1 (XCL1), C-C Motif Chemokine Ligands 8, 15, 21, 24, and 28 (CCL8, CCL15, CCL21, CCL24, CCL28), C-X-C Motif Chemokine Ligands 5 and 11 (CXCL5, CXCL11), Interferon Lambda 1 and 2 (IFNL1, IFNL2), Interleukin 24 and 34 (IL-24, IL-34), and Tumor Necrosis Factor Superfamily Member 10 (TNFSF10; TRAIL).

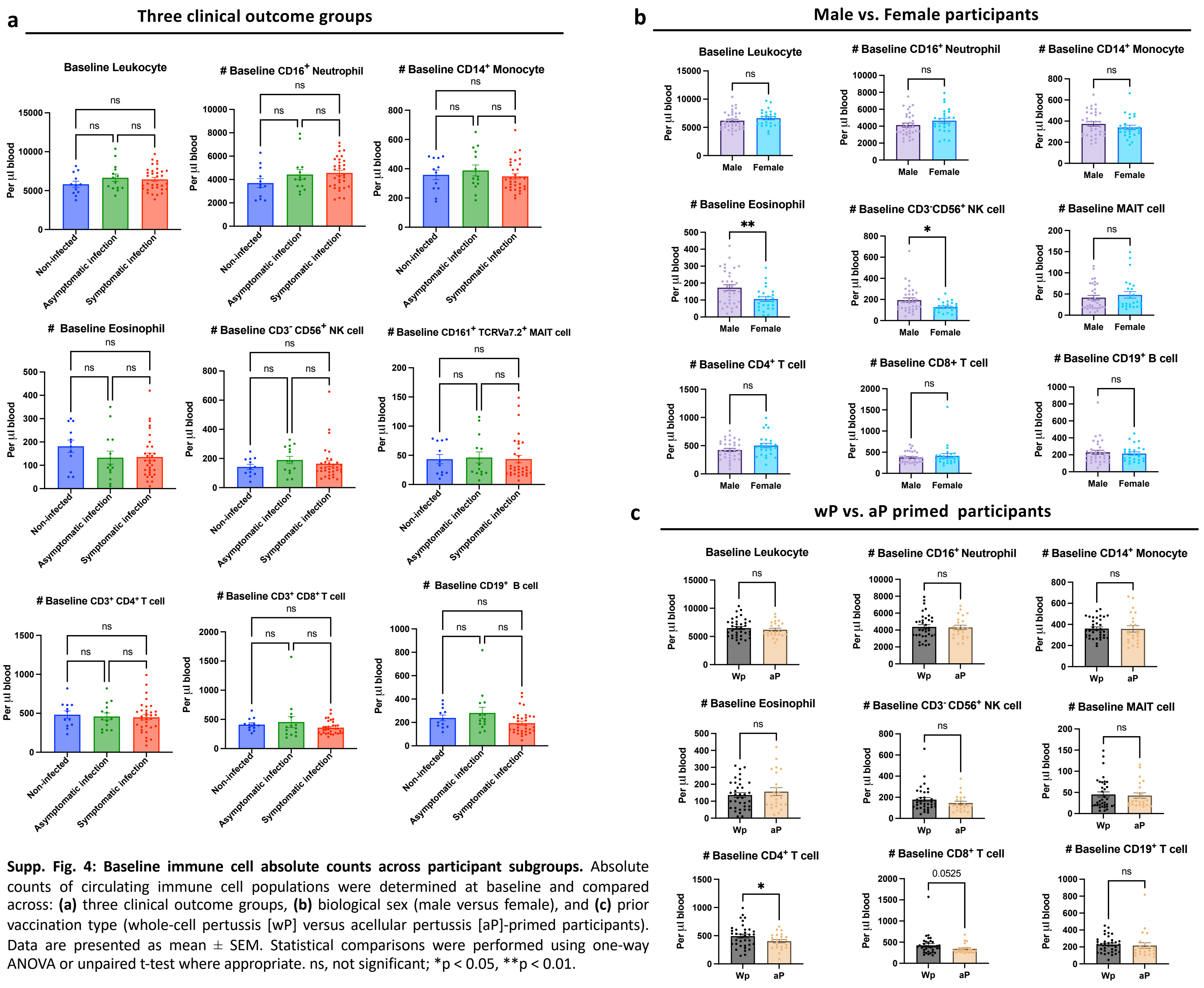


**Supp. Fig. 4: Baseline immune cell absolute counts across participant subgroups.** Absolute counts of circulating immune cell populations were determined at baseline and compared across: **(a)** three clinical outcome groups, **(b)** biological sex (male versus female), and **(c)** prior vaccination type (whole-cell pertussis [wP] versus acellular pertussis [aP]-primed participants). Data are presented as mean ± SEM. Statistical comparisons were performed using one-way ANOVA or unpaired t-test where appropriate. ns, not significant; $^{*}p < 0.05$, $^{**}p < 0.01$.

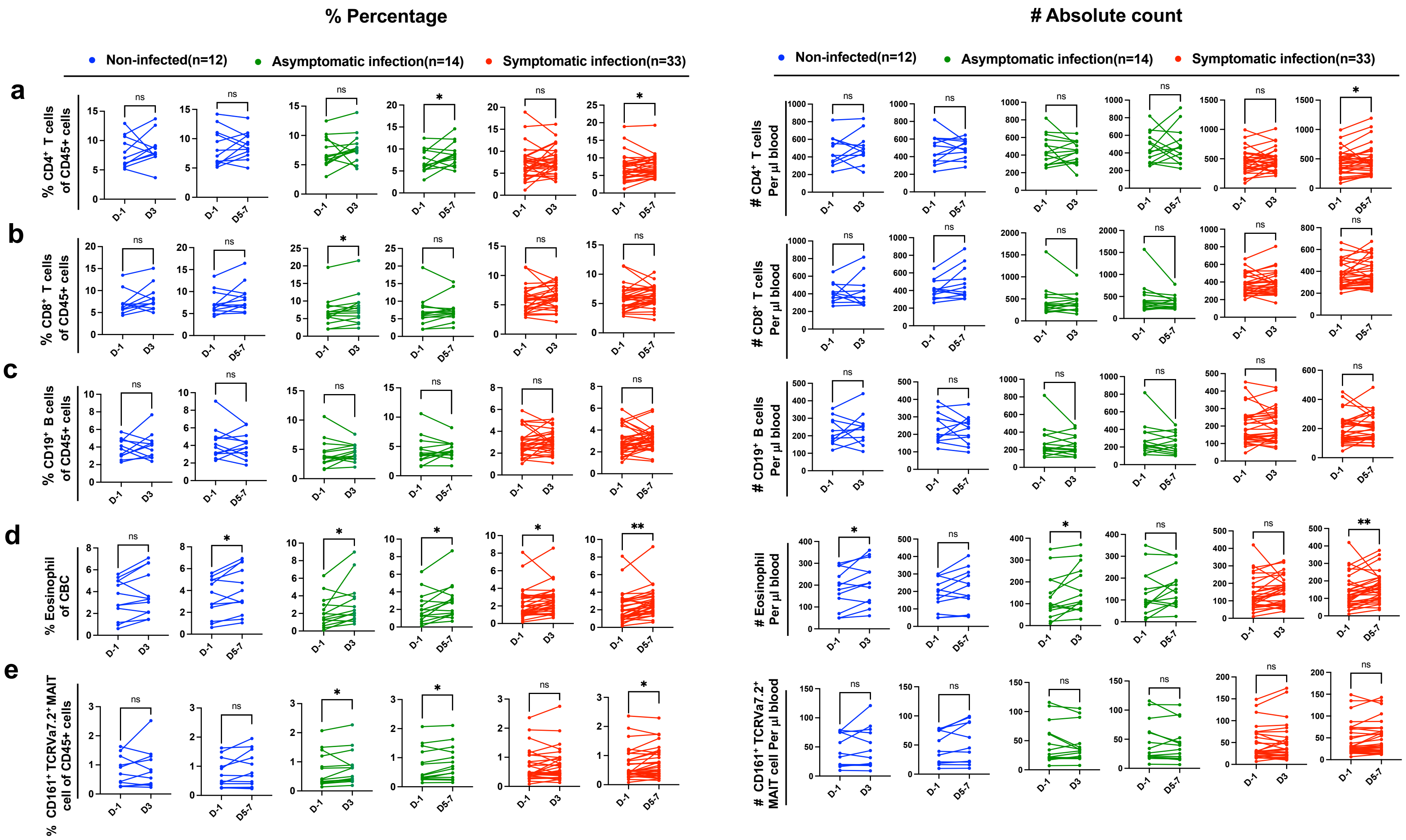


**Supp. Fig. 5: Adaptive and innate immune cell dynamic in peripheral blood following *B. pertussis* challenge.** Absolute cell counts and percentages measured at Day-1, Day 3, and mean of Day 5 and Day 7 post-challenge for: (a) CD3+CD4+ T cells, (b) CD3+CD8+ T cells, (c) CD19+ B cells, (d)Eosinophil (from CBC analysis), (e) MAIT cells. Data were tested for normality; normally distributed data were analyzed using a paired t-test, while non-normally distributed data were analyzed using the Wilcoxon signed-rank test. Dots represent all participant values. Statistical significance: ns, not significant; *p < 0.05, **p < 0.01.

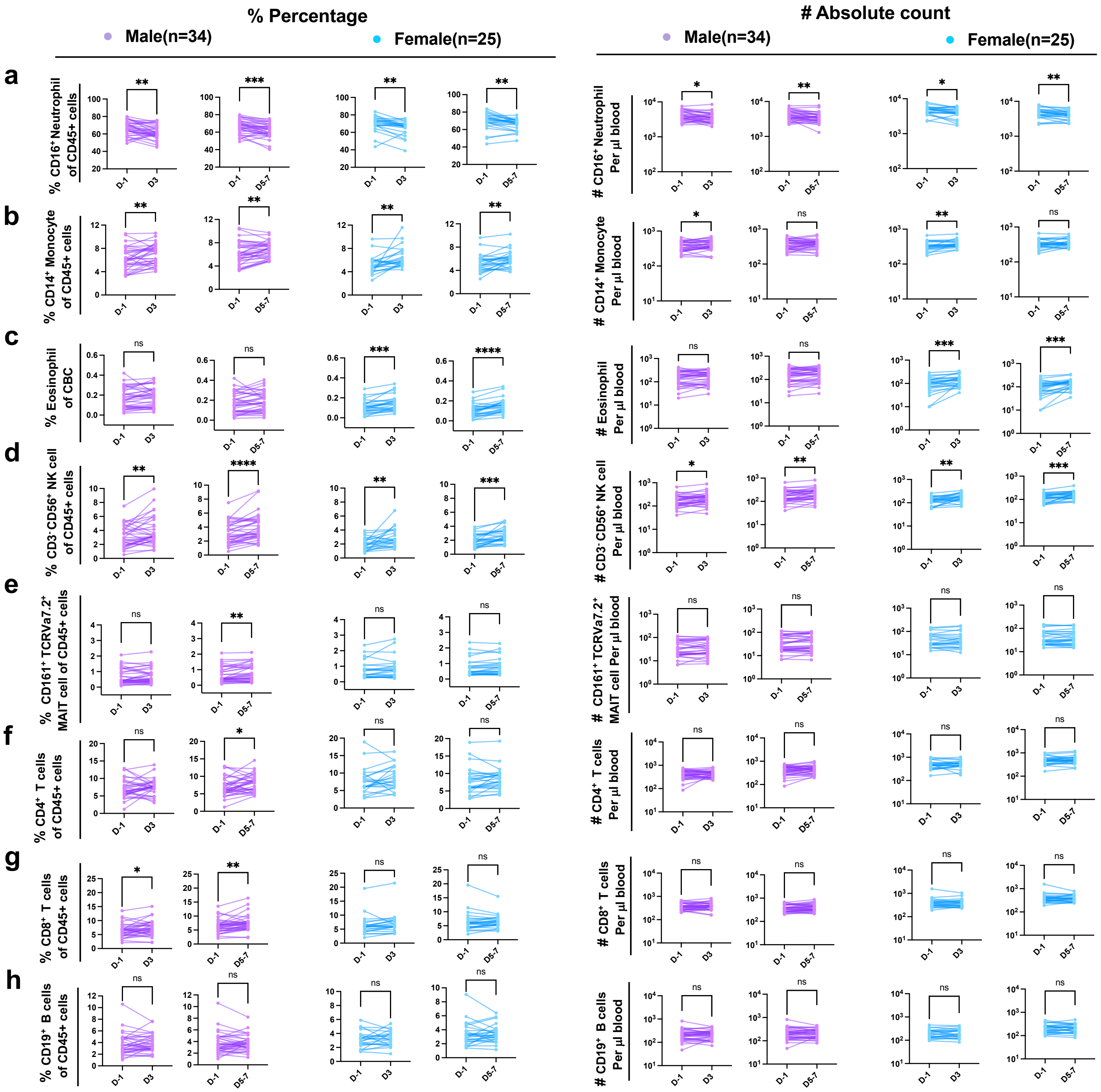


**Supp. Fig. 6: Immune cell dynamics in whole blood following *B. pertussis* challenge in female and male participants.** Absolute cell counts and percentages measured at Day −1, Day 3, and mean of Day 5 and Day 7 post-challenge by flow cytometry for: (a) $CD45^{+}CD16^{+}$ Neutrophils, (b) $CD45^{+}CD14^{+}$ Monocytes, (c) Eosinophils (from CBC analysis), (d) $CD45^{+}CD3^{-}CD56^{+}$ NK cells, and (e) $CD45^{+}CD3^{+}CD8^{+}CD161^{+}TCRV\alpha7.2^{+}$ MAIT cells, (f)CD3+CD4+ T cells, (g)CD3+CD8+ T cells , and (h) CD19+ B cells. Data were tested for normality; normally distributed data were analyzed using a paired t-test, while non-normally distributed data were analyzed using the Wilcoxon signed-rank test. Dots represent all participant values. Statistical significance: $*p < 0.05$, $**p < 0.01$, $***p < 0.001$, $****p < 0.0001$. CBC: Complete Blood Count.

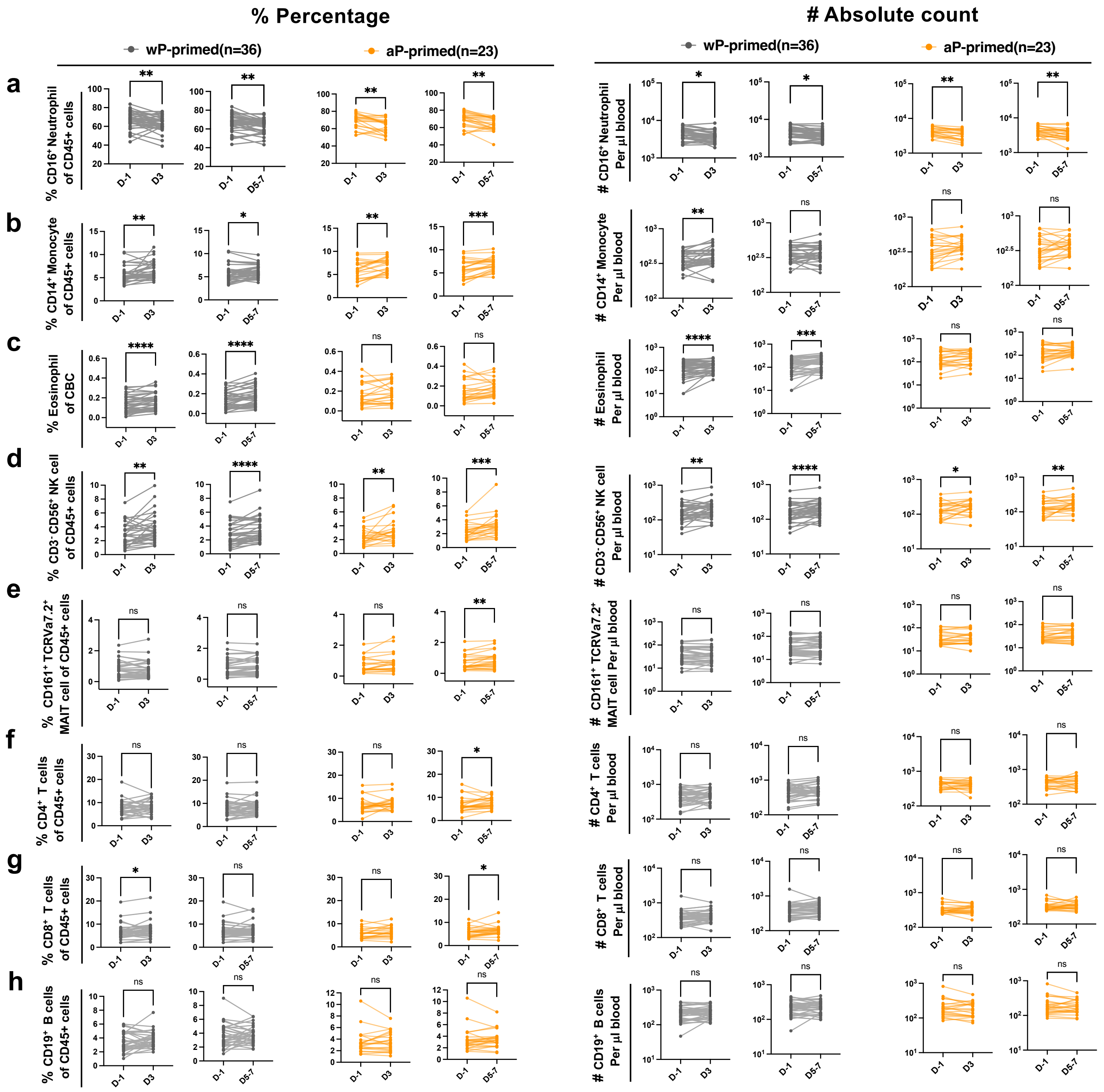


**Supp. Fig. 7: Immune cell dynamics in whole blood following *B. pertussis* challenge in participants with wP- or aP- vaccination history.** Absolute cell counts and percentages measured at Day −1, Day 3, and mean of Day 5 and Day 7 post-challenge by flow cytometry for: (a) $CD45^{+}CD16^{+}$ Neutrophils, (b) $CD45^{+}CD14^{+}$ Monocytes, (c) Eosinophils (from CBC analysis), (d) $CD45^{+}CD3^{-}CD56^{+}$ NK cells, (e) $CD45^{+}CD3^{+}CD8^{+}CD161^{+}TCRV\alpha7.2^{+}$ MAIT cells, (g)CD3+CD4+ T cells, (h)CD3+CD8+ T cells , and (h) CD19+ B cells. Data were tested for normality; normally distributed data were analyzed using a paired t-test, while non-normally distributed data were analyzed using the Wilcoxon signed-rank test. Dots represent all participant values. Statistical significance: *$p < 0.05$, **$p < 0.01$, ***$p < 0.001$, ****$p < 0.0001$. CBC: Complete Blood Count.

**Unstimulated NK cells**

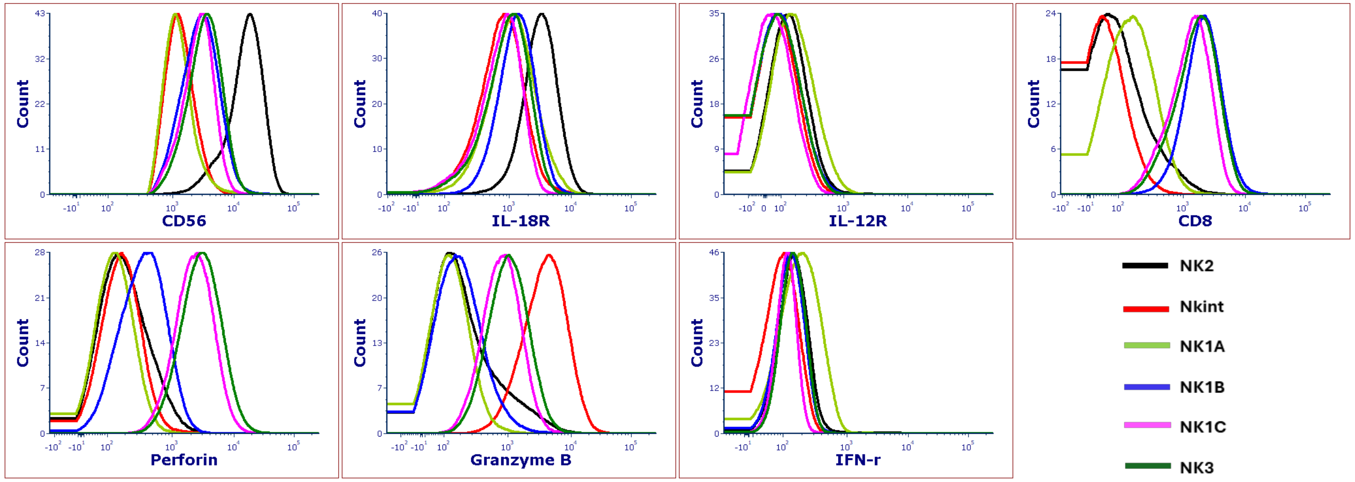


**HK *B. pertussis*-stimulated NK cells**

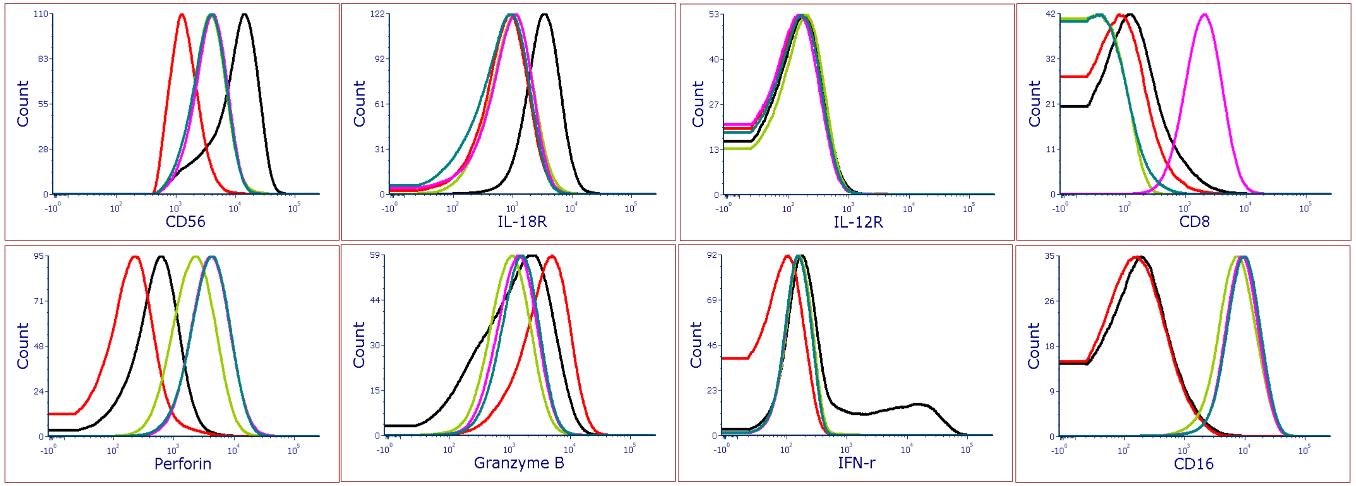


**Supp. Fig. 8: NK subset mapping.** PBMC samples cultured with or without HK *B. pertussis* stimulation were analyzed by flow cytometry. Six NK-cell subsets were identified on the t-SNE plot, and the expression profiles of surface markers and effector molecules were compared among subsets.

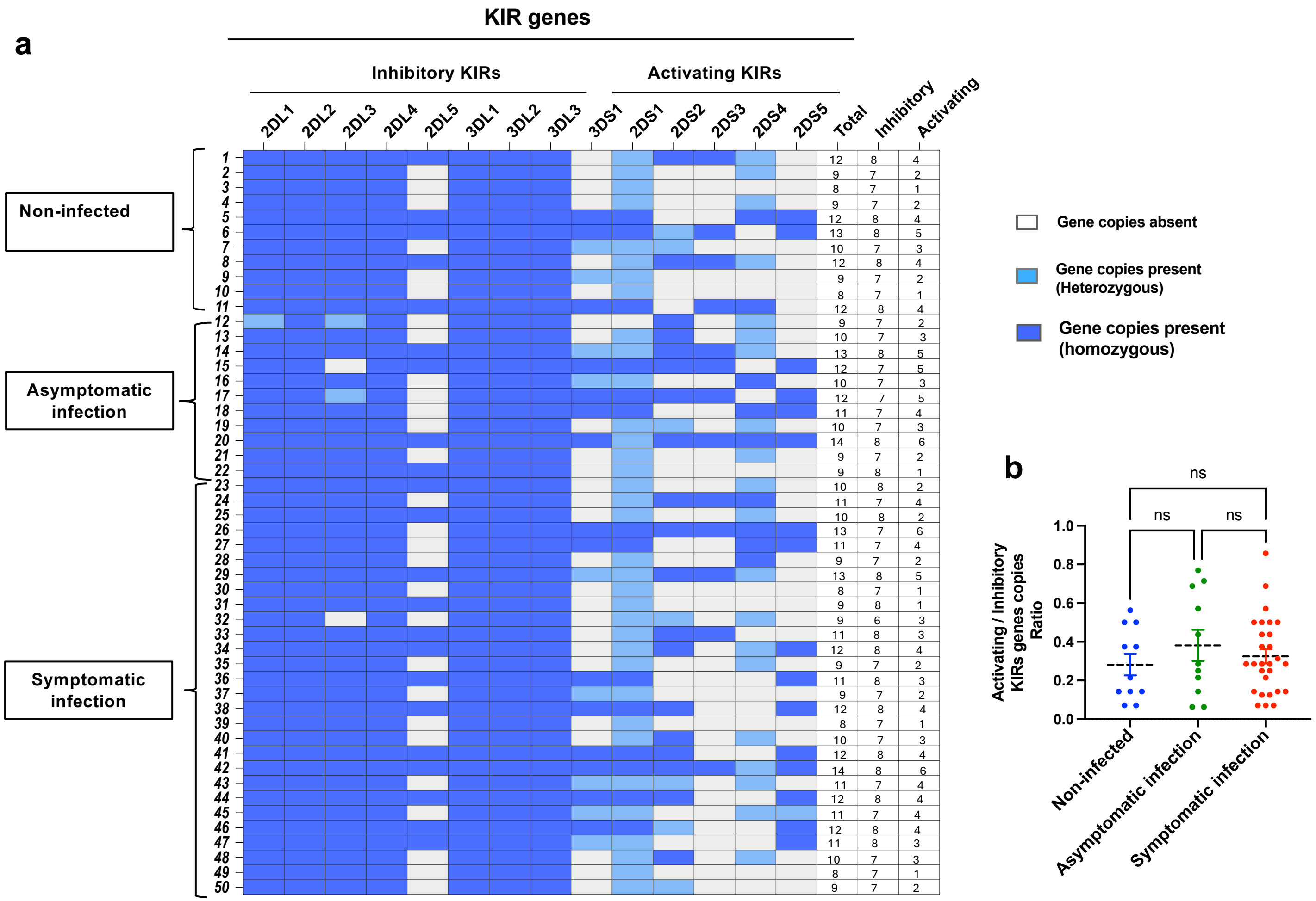


**Supp. Fig. 9: KIR genes presence/absence profiles and activating/inhibitory receptor distribution across individuals in different study groups. (a)** Heatmap showing the KIR gene repertoire of 50 individuals across three study groups: non-infected (individuals 1–11), asymptomatic infection (individuals 12–31), and symptomatic infection (individuals 32–50). Each row represents one individual, and each column represents a KIR gene, grouped into inhibitory KIRs (2DL1, 2DL2, 2DL3, 2DL4, 2DL5, 3DL1, 3DL2, 3DL3) and activating KIRs (3DS1, 2DS1, 2DS2, 2DS3, 2DS4, 2DS5). White cells indicate absence of gene copies; light blue indicates heterozygous gene copy presence; dark blue indicates homozygous gene copy presence. The total number of KIR genes, inhibitory KIR genes, and activating KIR genes per individual are shown in the three rightmost columns. **(b)** Dot plot showing the ratio of activating to inhibitory KIR gene copies for each individual across the three study groups (non-infected, asymptomatic infection, and symptomatic infection). Each dot represents one individual; Data are presented as mean ± SEM. Statistical comparisons were performed using one-way ANOVA. ns, not significant.

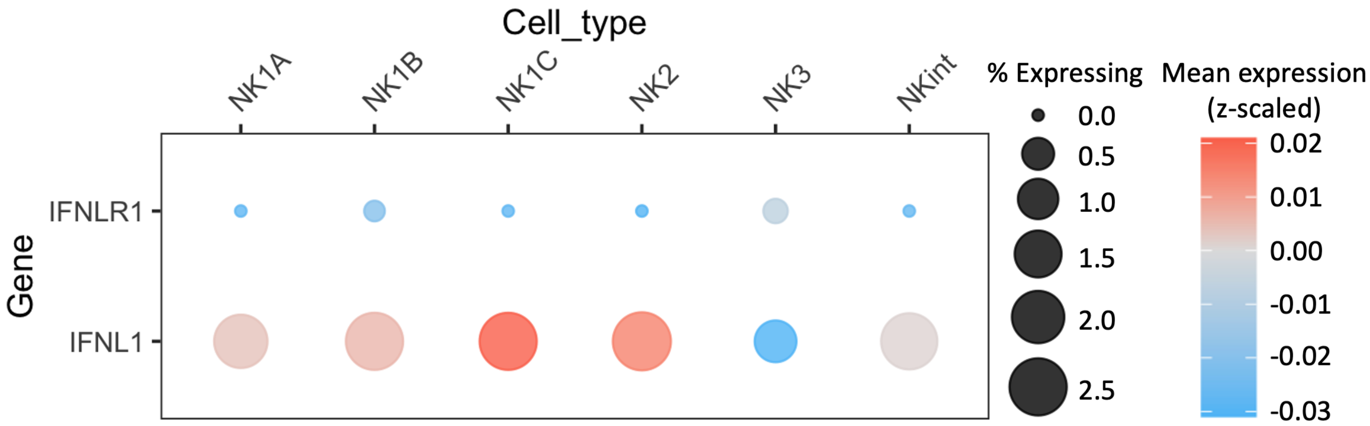


**Sup. Fig. 10: Transcriptomic analysis of IFNL1 (IL-29) and its receptor IFNLR1 by different subsets of NK cells. The data was obtained from the Human Cell Atlas and visualized as a heatmap in R.**

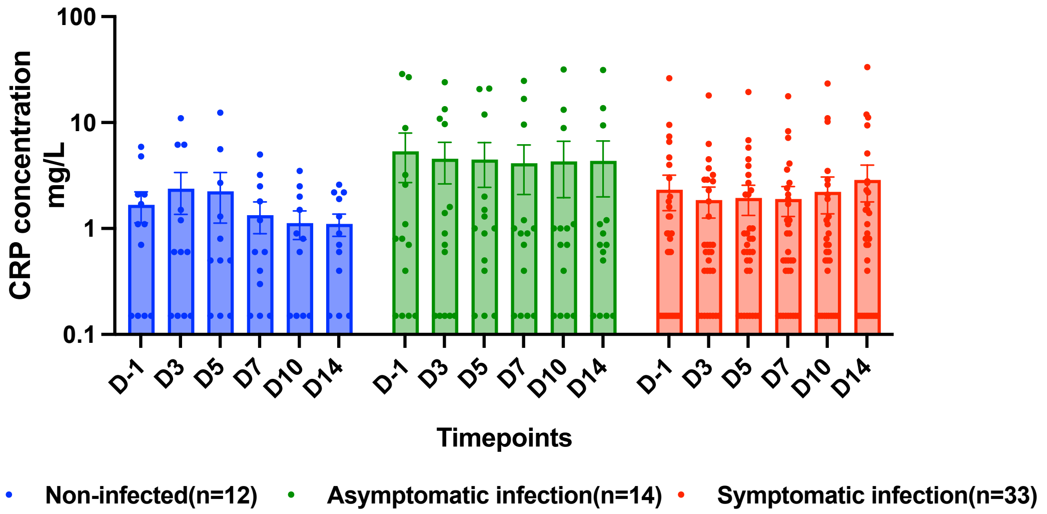


**Supp. Fig. 11: Longitudinal CRP concentration across study groups.** Serum C-reactive protein (CRP) concentrations (mg/L) were measured at six time points: baseline (D-1), D3, D5, D7, D10, and D14. Data are presented as mean ± SEM. Statistical comparisons were performed using two-way ANOVA followed by Bonferroni post test correction for multiple comparisons. A p-value of $<0.05$ was considered statistically significant.
